# Clinical-grade whole genome sequencing-based haplarithmisis enables all forms of preimplantation genetic testing

**DOI:** 10.1101/2023.12.06.23299605

**Authors:** Anouk E.J. Janssen, Rebekka M. Koeck, Rick Essers, Wanwisa van Dijk, Marion Drüsedau, Jeroen Meekels, Burcu Yaldiz, Maartje van de Vorst, Ping Cao, Bart de Koning, Debby M.E.I. Hellebrekers, Servi J.C. Stevens, Su Ming Sun, Malou Heijligers, Sonja A. de Munnik, Chris M.J. van Uum, Jelle Achten, Lars Hamers, Marjan Naghdi, Lisenka E.L.M. Vissers, Ron J.T. van Golde, Guido de Wert, Jos C.F.M. Dreesen, Christine de Die-Smulders, Edith Coonen, Han G. Brunner, Arthur van den Wijngaard, Aimee D.C. Paulussen, Masoud Zamani Esteki

**Affiliations:** Department of Clinical Genetics, Maastricht University Medical Centre (MUMC+), Maastricht, The Netherlands; Department of Genetics and Cell Biology, GROW school for Oncology and Reproduction, Maastricht University, Maastricht, The Netherlands; Faculty of Psychology and Neuroscience, Section Applied Social Psychology, Maastricht University, Maastricht, The Netherlands; Department of Human Genetics, Research Institute for Medical Innovation, Radboud University Medical Center, Nijmegen, The Netherlands; Department of Obstetrics and Gynaecology, GROW school for Oncology and Reproduction, Maastricht University, Maastricht, The Netherlands; Health, Ethics and Society, Maastricht University, Maastricht, Netherlands; GROW School for Oncology & Developmental Biology; CAPHRI School for Public Health & Primary Care, Maastricht University, Maastricht, The Netherlands; Division of Obstetrics and Gynaecology – Department of Clinical Science – Intervention & Technology (CLINTEC), Karolinska Institutet, Stockholm, Sweden

**Keywords:** pre-implantation genetic testing (PGT), whole genome sequencing (WGS), assisted reproductive technologies (ART), haplotyping, haplarithmisis

## Abstract

High-throughput sequencing technologies have increasingly led to discovery of disease-causing genetic variants, primarily in postnatal multi-cell DNA samples. However, applying these technologies to preimplantation genetic testing (PGT) in nuclear or mitochondrial DNA from single or few-cells biopsied from *in vitro* fertilised (IVF) embryos is challenging. PGT aims to select IVF embryos without genetic abnormalities. Although genotyping-by-sequencing (GBS)-based haplotyping methods enabled PGT for monogenic disorders (PGT-M), structural rearrangements (PGT-SR), and aneuploidies (PGT-A), they are labour intensive, only partially cover the genome and are troublesome for difficult loci and consanguineous couples. Here, we devised a simple, scalable and universal whole genome sequencing haplarithmisis-based approach enabling all forms of PGT in a single assay. In a comparison to state-of-the-art GBS-based PGT for nuclear DNA (37 embryos, 18 families, 25 indications), shallow sequencing-based PGT (10 embryos, 3 families), and PCR-based PGT for mitochondrial DNA (10 embryos, 2 families), our approach alleviates technical limitations by decreasing whole genome amplification artifacts by 68.4%, increasing breadth of coverage by 4-fold, and reducing wet-lab turn-around-time by 2.5-fold. Importantly, this method enables trio-based PGT-A for aneuploidy origin, an approach we coin PGT-AO, detects translocation breakpoints, and nuclear and mitochondrial single nucleotide variants and indels in base-resolution.

## Introduction

The Online Mendelian Inheritance in Man (OMIM) database lists over 4,700 genes with known phenotype-causing mutations^1^. Continuous advancements in sequencing technologies have substantially broadened our understanding of genetic disorders^2,3^, thereby increasing the potential indications for preimplantation genetic testing (PGT) for monogenic disorders (PGT-M). Several countries have initiated a population-wide offer of pre-conception carrier testing (PCT) for recessive disease, increasing the number of carrier couples^4–7^. In addition, continuous increases in maternal^8^ and paternal^9^ age, contribute to a higher risk for aneuploidies^10^, and *de novo* mutations^11^ and (segmental) chromosomal aberrations in offspring^12^, respectively. These demographic factors, together with the growing list of identified genetic diseases-causing mutations^13^, contribute to an increased demand for reproductive care, urging the need of scalable and generic genome-wide PGT approaches.

PGT is an assisted reproductive technology (ART), which is performed on DNA samples from one- or few-cell biopsies from day-3 or day-5/6 *in vitro* fertilised (IVF) embryos, respectively. PGT aims at selection of embryos unaffected from the genetic disorder in question or aneuploidies before intrauterine transfer, minimizing the need to contemplate pregnancy termination^14,15^. Since its inception in 1990^16^, PGT has evolved, encompassing non-hereditary genetic abnormalities i.e. aneuploidies, affecting embryo implantation and viability via PGT for aneuploidies (PGT-A). Yearly, 70,000 PGT cycles are performed globally^17–20^, with PGT-A representing over half of these in Euope^18^. PGT-A deems embryos harbouring any aneuploidies unsuitable for transfer. Despite randomised controlled trials (RCTs) casting doubt on the clinical utility of PGT-A^21–25^, this PGT approach remains popular. Furthermore, there is a growing body of literature demonstrating the birth of healthy, euploid children after the transfer of mosaic aneuploid embryos, as such the practise of discarding these embryos is increasingly criticised^26,27^. Existing whole genome sequencing PGT-A methods^28^, which rely on low-coverage embryo sequencing, lack parental information and do not allow determination of segregational origin of aneuploidies (*i.e.* whether it originates from meiosis or mitosis). PGT-M is the second most common form of PGT^18^ and traditionally required designing family- and locus-specific assays, tailored for the genetic disorder within the family, causing a long waiting list. While genome-wide haplotyping methods, such as Karyomapping^29^, haplarithmisis^30–32^, (S)Haploseek^33–35^, GENType^36^ provided a generic approach and alleviated this problem, these methods utilize SNP-genotypes of only a fraction of the genome and require complex laboratory and computational protocols. Currently, a substantial proportion of PGT-M procedures still rely on traditional approaches, such as PCR- or fluorescence in situ hybridisation (FISH)-based methods^18^. Comprehensive chromosome screening methods, such as VeriSeq, enable the assessment of chromosomal abnormalities in PGT for structural rearrangements (PGT-SR) with higher throughput than traditional methods including FISH^37^. Nonetheless, they cannot distinguish embryos with balanced translocations from those that are chromosomally normal^38^, nor can they detect haploid or triploid embryos. Importantly, transferring an embryo carrying a balanced translocation perpetuates the translocation, including the increased reproductive risk, to future generations. Another form of PGT, known as PGT for mitochondrial disorders (PGT-MT), focuses on mutations in the mitochondrial DNA (mtDNA) that contribute to the risk of inheriting a genetic mitochondrial disorder^39^. These mutations are exclusively maternally inherited and are characterized by heteroplasmy, defined as the coexistence of normal and mutated mtDNA. Clinical symptoms manifest only when the mutation load, the threshold of mutated mtDNA, surpasses a certain level. The mutation load inherited by offspring can be highly variable due to bottleneck principles. PGT-MT allows for selection of embryos carrying mutation loads below the pathogenic threshold^39^, minimizing the likelihood of clinical manifestation of the associated mtDNA disorder. The current prevailing approach for PGT-MT involves PCR-based method utilizing blastomere biopsy^39^. The integration of PGT-MT within the same laboratory workflow faces challenges due to limited availability of human data regarding the representativeness of the mutation load in TE biopsies for the entire embryo. All presented challenges underscore the need to develop a universal PGT method that streamlines laboratory protocols and provides comprehensive genome-wide coverage to address genetic disorders, even in complex genomic loci.

We devised a simplified, scalable, and universal whole genome sequencing-based method for PGT (WGS-PGT) that enables all forms of PGT within a single assay. Here, we demonstrate that WGS-PGT enables (i) PGT for genetic indications in complex genomic regions, (ii) direct detection of single- and few-base pair genetic variations, (iii) a novel form of PGT-A that uncovers segregational origin (meiotic vs. mitotic) of aneuploidies and their level of mosaicism, called PGT-AO, (iv) (in)direct detection of the translocation breakpoints and inheritance of normal and derivative chromosomes, allowing the distinction between normal embryos and balanced translocation carriers, and (v) PGT for mtDNA disorders.

## Results

### Proof-of-concept for WGS-PGT

To establish WGS for haplarithmisis-based PGT, we performed a pilot study in which we carried out deep sequencing (30-40X) to compare current clinical gold standard GBS-PGT (**Fig. 1a**) in two PGT families (family 1 and 2 with *n* = 4 embryos and *n* = 2 embryos, respectively) (**Fig. 1b**). The WGS method allows for a 2.5-fold reduction in library preparation times compared to GBS (**Fig. 1c**). We then performed *in silico* subsampling at target coverages of 5X, 10X, 20X and 30X, to determine the optimal depth of coverage for accurate diagnosis. To this end, we evaluated several key parameters, including breadth (**Fig. 1d**) and depth of coverage (**Supplementary** Fig. 1), Mendelian inconsistency rates (**Supplementary** Fig. 2) and haplotype concordance (**Supplementary** Fig. 3). WGS provided a 4-fold higher breadth of coverage, i.e. the proportion of the genome that is sequenced, than GBS, with WGS exhibiting a breadth of coverage exceeding 80% while GBS remains below 20%. Comparing different depth of coverage levels revealed a significant difference in breadth of coverage between 5X and 10X (*P* = 3.52 × 10^-3^, two-sided Wilcoxon’s rank-sum), while increasing the depth of coverage from 10X to 20X did not lead to a significant increase in breadth of coverage (*P* = 8.31 × 10^-2^, two-sided Wilcoxon’s rank-sum, **Fig. 1d**). The number of genome-wide informative SNPs increased 10-fold for WGS-PGT at 10X coverage compared to GBS-PGT, reaching 2.5 million genome-wide informative SNPs (± 46,000 s.d.) (**Supplementary** Fig. 4a). Specifically, at 5X coverage there were 1.5 million genome-wide informative SNPs (± 53,422 s.d.), whereas at 30X coverage, this number increased to 2.9 million (± 25,960 s.d.). Typically, a higher number of genome-wide informative SNPs increases the accuracy and reliability of haplotype inference. For all target coverages, the mean genome-wide haplotype concordance between GBS and WGS was higher than 97%. Increasing the depth of coverage from 10X to 20X, revealed no significant improvement (paternal haplotype *P* = 0.59 and maternal haplotype *P* = 0.70, two-sided Wilcoxon’s rank-sum, **Supplementary** Fig. 3). Furthermore, the mean Mendelian inconsistency rates representing WGA artefacts reduced substantially from 11.3% (± 1.26 s.d.) in GBS-PGT to 4.6% (± 1.35 s.d.) for WGS-PGT at 10X coverage (**Supplementary** Fig. 2a). Based on these findings, we settled that 10X sequencing provided sufficient data to reliably conduct haplarithmisis-based PGT-M.

**Fig. 1:**
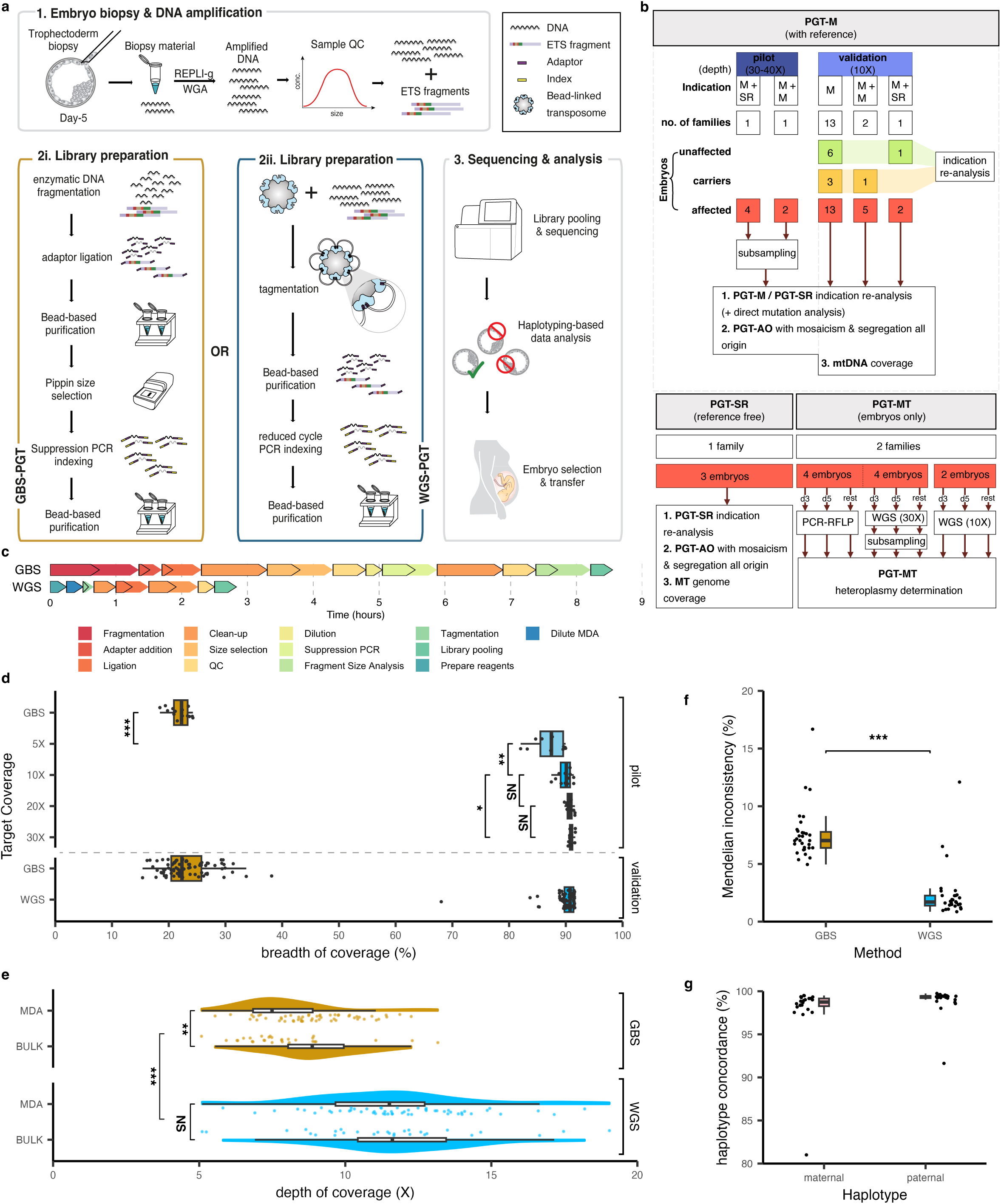
Overview study workflow and key parameters. **a,** The study’s workflow including embryo biopsy, DNA amplification, library preparation (GBS and WGS), sequencing, analysis, and clinical diagnosis. **b,** study sample inclusion diagram. **c,** library preparation times in hours for GBS and WGS methods. Black outline represents hands-on time per step. **d,** boxplot showing the breadth of coverage for subsampled WGS samples (*n* = 12 per target coverage) compared to corresponding GBS samples and for 10X WGS samples (*n* = 84). **e,** half violin plot, overlayed with boxplot showing depth of coverage for WGAed embryo and bulk gDNA samples for GBS and WGS. Each dot represents an individual sample. **f,** Mendelian inconsistency rate for 10X embryo samples (*n* = 31 per method). **G,** genome-wide haplotype concordance of WGS with GBS indicated per parent (maternal, *n* = 23; paternal, *n* = 25). In all boxplots the horizontal lines of the boxplot represent the 25^th^ percentile, median and 75^th^ percentile. The whiskers extend to 1.5 * IQR. GBS: genotyping-by-sequencing; WGS: whole genome sequencing; IQR: interquartile range; WGAed: whole genome amplified.

### Clinical validation of WGS-PGT

To clinically validate WGS-PGT, we sequenced 31 embryo samples from 16 families at 10X coverage (**Fig. 1b, Supplementary Table 1**). Specifically, we selected PGT families that posed analytical challenges when using GBS-PGT e.g. when the region of interest (ROI), representing the genomic location of the pathogenic variant of interest, was located in telomere regions, the family was consanguineous, the family had multiple genetic disorders, such as monogenic diseases and translocations or when the embryo exhibited a haplotype recombination in proximity of the ROI. We observed a disparity in depth of coverage between Whole Genome Amplified (WGAed) embryo and bulk samples for GBS (*P* = 1.43 × 10^-3^, two-sided Wilcoxon’s rank-sum), which is likely attributed to the potential loss of restriction enzyme sites during amplification, leading to a lower library amount (**Fig. 1e**). However, in WGS, we found a similar depth of coverage between bulk DNA and WGAed samples, indicating a more stable read out from WGS-PGT (*P* = 0.46, two-sided Wilcoxon’s rank-sum). Results from the key parameters could be replicated in these families, with a mean autosomal Mendelian inconsistency rate of 2.42% (± 2.23 s.d.) for WGS and mean Mendelian inconsistency of 7.66% (± 2.33 s.d.) for GBS (*n* = 31 embryos) (**Fig. 1f**). The mean autosomal Mendelian inconsistency rate in the validation subset demonstrated a lower rate than in the pilot study. This difference can be attributed to the specific characteristics of family 1, where all embryos had a translocation and two embryos harboured an aneuploid chromosome (**Supplementary** Fig. 2a**, Supplementary Table 4**). Moreover, the mean concordance of maternal haplotypes was 97.8% (± 3.9 s.d.), and mean concordance of paternal haplotypes was 99.0% (± 1.6 s.d.) (**Fig. 1g**). One outlier was observed at 81.0% for maternal haplotype concordance, originating from an embryo with a triploid genome. One of the diagnostic criteria in the analysis of an embryo for a specific monogenic indication include the number of informative SNPs in a 4 Mb interval, *i.e.* 2 Mb up- and downstream of the mutation, are considered on the maternally and/or paternally inherited haplotypes if these meet our assessment criteria (**Methods** and **Supplementary Table 2**). Since we have deliberately selected challenging PGT families with ROI in complex genomic regions or high rates of consanguinity, in 13 out of 35 ROI the haplotype concordance did not meet the assessment criteria in GBS-PGT (**Supplementary Table 3**). However, using WGS-PGT, in 5 out of the 13 ROIs, the assessment criteria could be met, owing to its inherent higher resolution.

### PGT-M with the potential to directly detect pathogenic single nucleotide variants

PGT-M is challenging when a close relative is unavailable for phasing, or in cases when prospective parents present with a *de novo* pathogenic single nucleotide variant (SNV). Detection of the pathogenic variant shows a promising alternative as it may facilitate a diagnosis in these families. While GBS approaches only cover 20% of the genome and enabled indirect detection monogenic aberrations, WGS-PGT, at 10X depth of coverage, covers >80% of the genome (**Fig. 1d**), thereby facilitating direct SNV detection. For 22 monogenic indications that included single base pair substitutions or deletions, we compared the genotypes and diagnoses ascertained from direct SNV detection with those anticipated based on the haplarithmisis result. Direct SNV detection provided the correct diagnosis in 90% (*n* = 20) and correct genotype in 82% (*n* = 18) of the ROI (**Fig. 2b, Supplementary** Fig. 6). When the number of reads at the ROI was higher than 5, the expected genotype could be correctly identified in all cases. Remarkably, direct variant detection showed promise in rare instances, resolving pathogenic SNVs in embryos with inconclusive haplarithmisis results. Specifically, in embryo 23, which was assessed for an autosomal dominant pathogenic SNV, the assessment criteria thresholds for haplarithmisis were not met (**Supplementary Table 2**), resulting an inconclusive diagnosis of the embryo. Direct variant detection showed presence of the mutant allele at the ROI in 4 out of 5 reads, allowing embryo 23 to be classified as affected (**Fig. 2b, Supplementary** Fig. 6). To examine whether direct pathogenic variant detection could also be applied to larger deletions, we visualised indications representing deletions of two or more base pairs (*n* = 11). Deletions spanning two or three base pairs could be identified within the integrated genomics viewer software (IGV) (*n* = 6, **Supplementary** Fig. 7a). Bigger deletions presenting as autosomal recessive pathogenic variants showed a loss of coverage (*n* = 2, **Supplementary** Fig. 7b), while bigger deletions presenting as autosomal dominant or x-linked variants could be identified using the “view as pairs” option in IGV in two affected embryos from family 7 and two unaffected embryos could be confirmed (**Supplementary** Fig. 7c). Furthermore, one aberrant embryo did not show the mutation as it has a mitotic trisomy with of the unaffected haplotype. Notably, the putative deletion could not be detected directly in one embryo (embryo 5 from family 2) which was a carrier of the deletion (**Supplementary Fig.7c**).

**Fig. 2:**
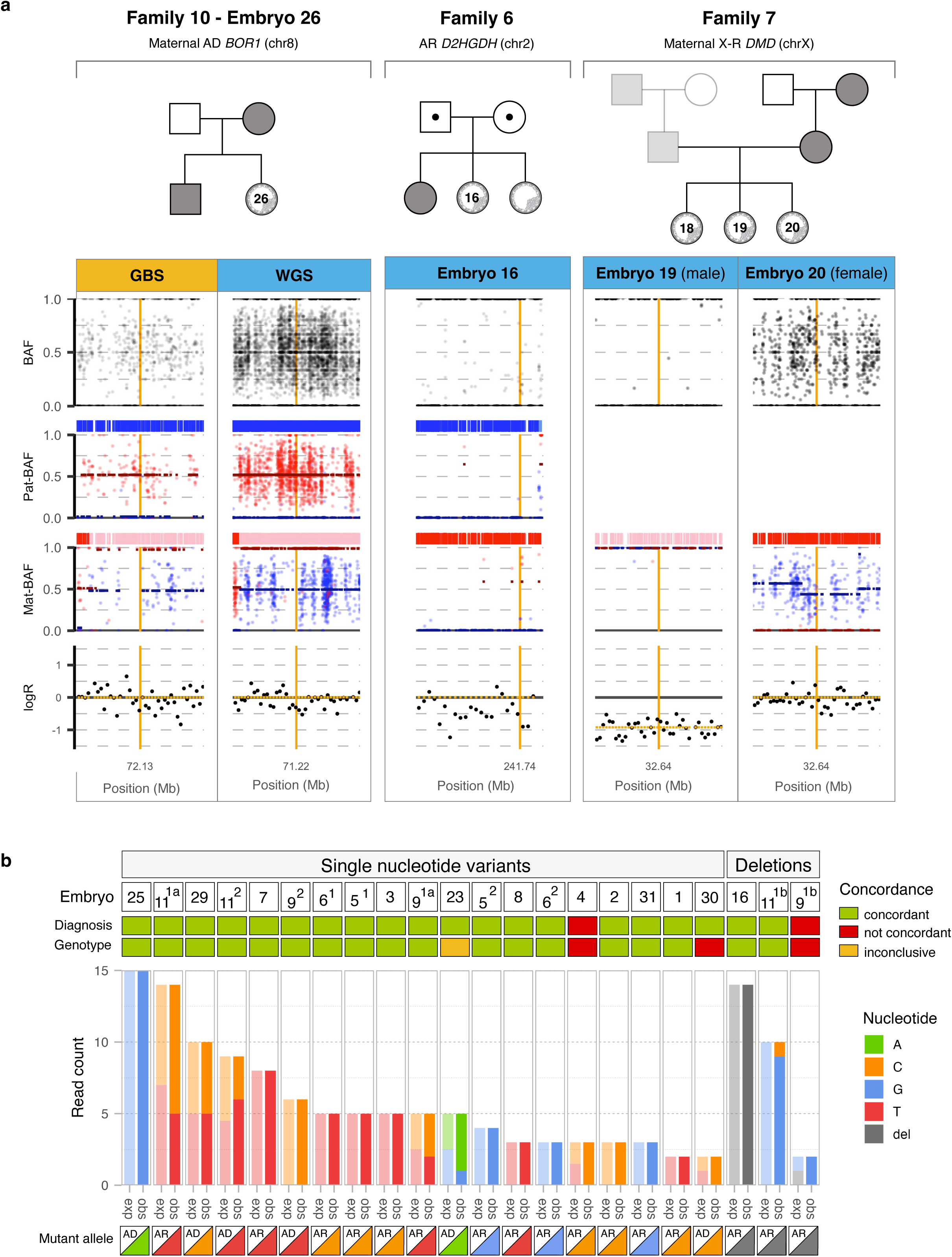
PGT for monogenic disorders. **a,** Representative haplarithms for all types of PGT-M indications, *i.e.* AD (family 10), AR (family 6), X-R (family 7), and highlighting the increased resolution of WGS-PGT compared to GBS-PGT. The top panel shows family pedigrees. The lower panel shows haplarithms generated with GBS (gold) and WGS (blue) – from top to bottom – BAF profiles, interpreted paternal haplotypes (paternal H1: darkblue; paternal H2: lightblue), Pat-BAFs, interpreted maternal haplotypes (maternal H1: red; maternal H2: pink), Mat-BAFs, logR values representing the log2 ratio of the observed copy numbers to the expected copy number. The ROI is indicated by an orange line. **b,** Genotypes ascertained by direct mutation detection (obs) compared to the haplarithmisis result (exp) subdivided for genetic indications that included SNVs or single nucleotide deletions. Embryos are numbered, and the indications are indicated in superscript (1: indication 1, 2: indication 2, a or b indicate different genetic loci in case of a compound heterozygous mutation). The mutant allele track shows the type of inheritance indicated by AR or AD, and the triangle is coloured with the mutant allele (A = green, C = orange, G = blue, T = red, del = grey). The concordance of diagnosis and genotype with the haplarithmisis result is colour-coded (concordant = green, not concordant = red, inconclusive = orange). Recessive indications were considered inconclusive if the proportion of reads supporting the reference allele were <30% or >60%. PGT: preimplantation genetic testing; M: monogenic; AR: autosomal recessive; AD: autosomal dominant; X-R: X-linked recessive; BAF: B-allele frequency; Pat: paternal; Mat: maternal; ROI: region of interest; GBS: genotyping-by-sequencing; WGS: whole genome sequencing; exp: expected, obs: observed; del: deletion.

### PGT for aneuploidy origin (PGT-AO): a transformative PGT-A

Haplarithmisis-based WGS-PGT can accurately determine the segregational origin, *i.e.* meiosis I, meiosis II, or mitosis, of aneuploidies and their degree of mosaicism (>10%) (**Fig. 3a, Supplementary Note** **Fig. 3a**). Unlike meiotic trisomies, which involve both homologous chromosomes of the contributing parent, mitotic trisomies result from the exact duplication of a single homologue. Distinction between mitotic and meiotic II trisomy is possible when a crossover takes place on the chromosome of interest. The crossover rate for human chromosomes varies between 1.07 cM/Mb to 1.76 cM/Mb, ensuring that each chromosome generally experiences at least one crossover, while the actual number of crossovers depends on the crossover rate and chromosome size^40^. Distinction between mitotic and meiotic II trisomies is not possible in rare cases where there is no crossover, or when the crossover is located in a challenging genomic region (e.g. telomeric or centromeric).

**Fig. 3.**
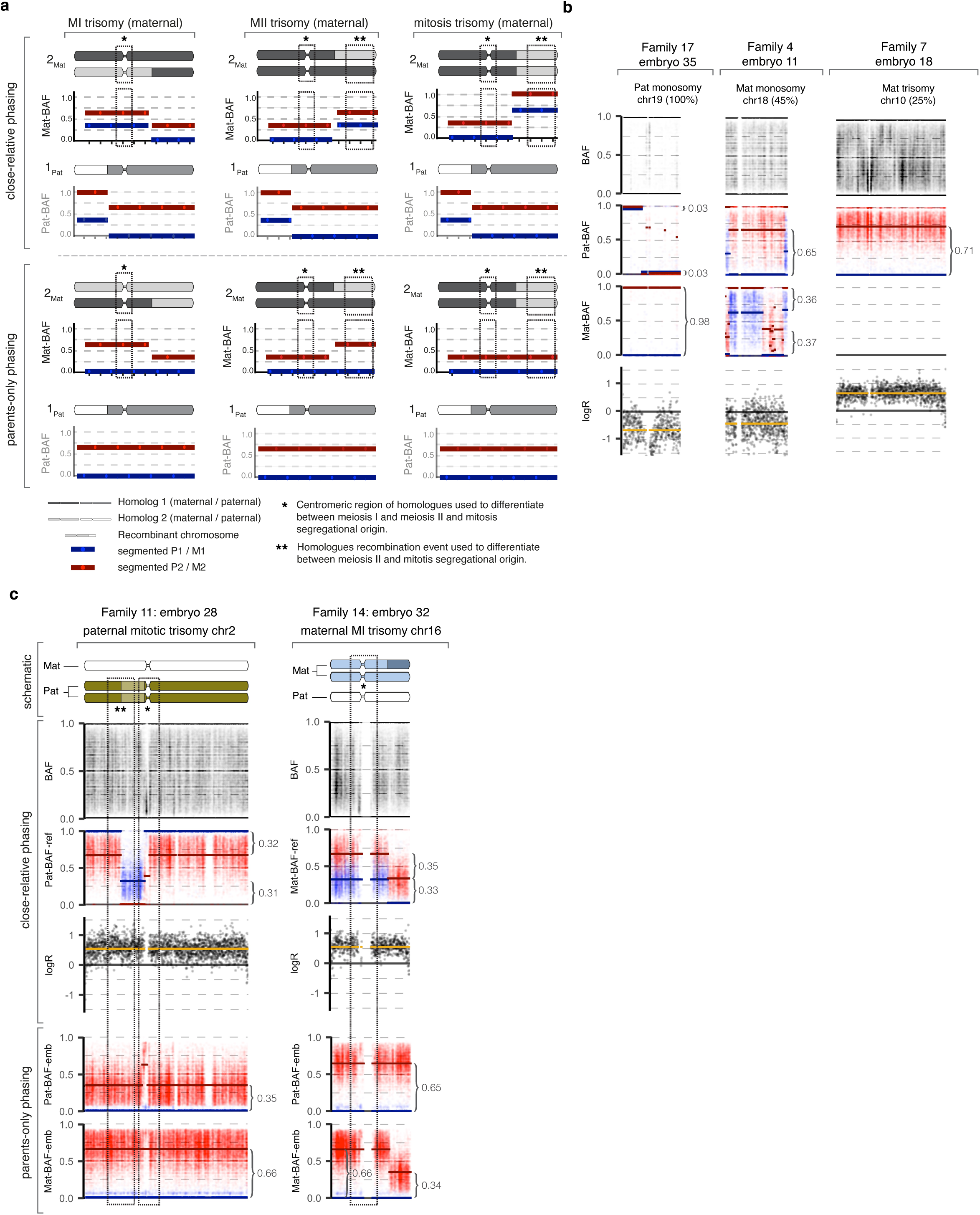
PGT for aneuploidy origin. **a,** Schematic representation of trisomies with different segregational origins (meiosis I, meiosis II or mitotic trisomy) are shown, using close-relative phasing or using parents-only phasing to identify segregational origin. The segregational origin using parents-only phasing was determined by inspecting the distance between the segmented P1/M1 and P2/M2 at the centromeric and telomeric region. Further explanation is provided in Supplementary Note. **b,** Representative cases of aneuploidies with different degrees of mosaicism. The degree of mosaicism was determined by calculating the distance between paternal and maternal BAF segments as indicated with curly brackets. **c,** Representative haplarithms of embryos with trisomies with a different segregational origins. Top panel: schematic of the chromosomal constitution. Middle panel: haplarithms when using a close relative as referent. Bottom panel: haplarithms when using the same embryo as referent to phase the parents and define the segregational origin. Curly brackets indicate the distance between the paternal and maternal BAF segments.

We identified 14 aneuploidies in 11 of 29 affected embryos analysed, encompassing 10 trisomies, 3 monosomies and 1 triploidy, with some embryos carrying more than one aberration. The segregational origin could be determined for 10 out of 11 detected chromosomal gains (10 trisomies and 1 triploidy, **Supplementary Table 4**). Only a single embryo (**Fig. 3b** - embryo 18, family 7) did not have a crossover in the trisomic chromosome, precluding the differentiation between meiotic II and mitotic origins. Furthermore, we observed mosaic meiotic trisomies suggesting that a fraction of the biopsied cells had undergone chromosomal rescue (**Supplementary Table 4**)^28^. Three mosaic aberrations with a meiotic origin were identified (**Supplementary Table 4**). These included two embryos with a trisomy with a mosaicism level of 80% and 90%. The third embryo had a genome-wide triploidy with meiosis II origin and mosaicism level of 100% while chr15 was diploid with a mosaicism level of 50%, suggesting that a fraction of the biopsied cells underwent chromosomal rescue for chr15^41^. Additionally, the mosaicism level for 3 embryos with monosomy was determined (100%, 100%, 45%). Mosaicism levels of 0% and 100% indicate a non-mosaic zygosity. We further assessed nine aberrant embryos with copy number gains of known segregational origin to validate parents-only haplotyping and subsequent determination of segregational origin (**Fig. 3c**). In all cases the segregational origin detected by parents-only haplarithmisis was concordant with standard haplarithmisis. These results expand the possibilities of detecting segregational origin in human preimplantation embryos when no close relative is available.

### PGT-SR with direct and indirect detection of translocation breakpoints

Nine embryos from three families that underwent PGT-SR with shallow sequencing (**Methods**) were re-analysed using the WGS-PGT protocol. The copy number state of all embryos could be correctly determined with WGS-PGT and haplarithmisis purely by assessing the segmentation of the logR values, which represent the log2 ratio of the observed to expected copy number (**Fig. 4, Supplementary** Fig. 8**, Supplementary Table 5**). Although small duplications, such as the 1.08 Mb segmental duplication of chr16 in embryo 4 of family 1, were not segmented, such embryos could still be correctly diagnosed based on the presence of the reciprocal deletion (**Fig. 4**). The identification of these segmental deletions and duplications remained consistent across all subsampled sequencing depths, even at 5X coverage (**Supplementary** Fig. 8). In couples where embryos exhibited no copy-number imbalances, the possibility of inheriting either both normal homologues or both derivative chromosomes (balanced translocation) from the carrier parent should be considered. Shallow sequencing proves insufficient to distinguish between these cases. However, our haplarithmisis-based PGT successfully identified embryos with unbalanced translocations. For these cases, the diploid flanking haplotypes of the translocation allowed us to distinguish between normal and derivative chromosomes in all other embryos. Specifically, embryo 1 and 4 from family 1 were phased with an unaffected sibling as a seed for phasing. Haplotypes on the diploid side of the chromosomal breakpoint were distinguished as either dark blue, indicating consistency with the reference haplotype or light blue, indicating the alternative haplotype. Consequently, embryo 1 inherited the normal chr8 and derivative chr16 while embryo 4 inherited the derivative chr8 and normal chr16 (**Fig. 4**). Importantly, the paired-end sequencing data of the carrier parent and the embryos can be leveraged to identify translocation breakpoints. In all carrier parents (*n* = 3) included in this study, we could identify a breakpoint pair that closely corresponded to the expected translocation breakpoints ascertained using karyotyping (**Supplementary Table 5,** sheet 1-3). Unlike the karyotyping results, the breakpoints derived from paired-end sequencing data could be determined at approximately base pair resolution, allowing a more precise regions of interest to be defined for haplarithmisis. It is important to note that the breakpoint locations obtained may exhibit variation among embryos, and in some cases, multiple hits may be identified. Subsampled data from family 1 showed that a minimum genome-wide depth of coverage of 10X is required to accurately call these breakpoints (**Supplementary Table 5,** sheet 1). We could identify corresponding breakpoint pairs in six of the eight unbalanced embryos from families 1 and 19. The two embryos in which no breakpoints were identified both inherited a derivative chr8 carrying the small segment of chr16 (1.08Mb). Although paired-end sequencing analysis did not identify a breakpoint pair in embryo 23 of family 9, which could be attributable to a chaotic copy number profile (**Supplementary Table 5,** sheet 4), we could correctly identify the relevant breakpoint pair in the unbalanced embryos 24 and 25. For the paternal ins(10;7) of family 19 (**Supplementary Table 5, sheet 2**), one would expect to find a single breakpoint on chr10 and two corresponding breakpoints on chr7, specifically a position on the q arm of chr7 and the end of chr7. However, this could not accurately be detected using Manta. While Manta did identify breakpoint pairs from between chr10 and chr7 in the carrier parent, the chr10 coordinates do not match the findings from the diagnostic karyotyping and only one corresponding chr7 position was identified which lies in the centre of the expected inserted segment. Whether the same breakpoints would be found in unbalanced embryos could not be assessed as neither of the unbalanced embryos, embryos 38 and 39, inherited the derivative chr10. Based on the segmented logR information from Haplarithmisis conducted on embryos 38 and 39, it was however possible to determine that the chr7 breakpoint would be around 129,135,000 bp, which is in keeping with the findings from the diagnostic karyotyping of the father.

**Fig. 4.**
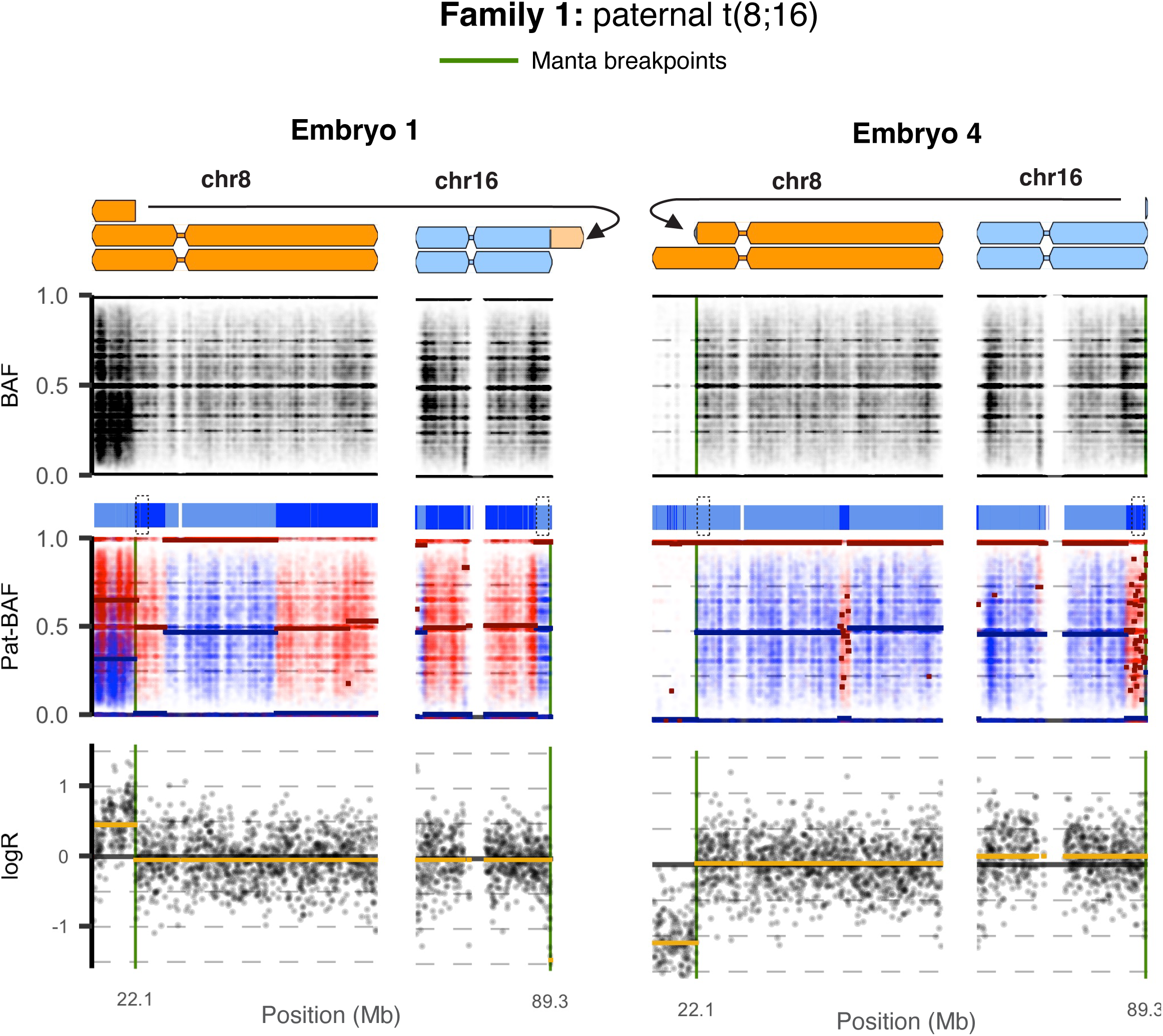
PGT for structural rearrangements. Representative haplarithms for embryos with structural rearrangements. Two embryos from family 1 are shown where the structural rearrangement is a translocation between chr8 and chr16 as indicated by the schematic chromosome representation. Haplarithms include BAFs, paternal haplotypes, Pat-BAF and copy number indicated by logR with breakpoints as identified by Manta (green) and flanking haplotypes H1 (darkblue) and H2 (lightblue) indicated by the dashed box. PGT: preimplantation genetic testing; BAFs: B-allele frequencies; Pat: paternal; H1: haplotype 1; H2: haplotype 2.

### PGT for mitochondrial DNA disorders

The current gold standard for PGT for mtDNA disorders (PGT-MT) requires a specialised PCR-based restriction fragment length polymorphism (PCR-RFLP) workflow that is carried out on a day-3 blastomere biopsy^39^. We compared the heteroplasmy levels from the blastomere biopsy to heteroplasmy levels obtained by applying the same protocol to day-5 trophectoderm (TE) biopsies (*n* = 4, **Fig. 1b**) and found that the heteroplasmy levels differed by 1% to 4% (**Fig. 5a**). Furthermore, the PCR-RFLP protocol applied to the corresponding DNA derived from the surplus embryos, i.e. the remaining embryo, yielded heteroplasmy levels that differed by 0% to 4% and 1% to 3% to the day-3 and day-5 biopsy results, respectively (**Fig. 5a**). Subsequently, we applied our WGS-PGT protocol to day-5 TE biopsies (*n* = 4), and DNA derived from the corresponding surplus embryo material, *i.e.* 100-200 cells, with a target genome-wide sequencing depth of 30X (**Fig. 1b**). The mitochondrial genome was highly covered at all levels of subsampling with only one site being covered less than 100 times in the TE-biopsy data after subsampling to 10X coverage (**Fig. 5b**). Importantly, 105 mtDNA sites with known pathogenic variants had a minimum coverage of 1,944X in the 10X subsampled TE-biopsy data (*n* = 4) (**Supplementary** Fig. 9). To assess reproducibility of our PGT-MT approach, we calculated the mitochondrial genome coverage of the samples sequenced at 10X for PGT-MT (*n* = 2) or PGT-M and PGT-SR (*n* = 23) indications and found that the coverage across all WGAed embryo samples was comparable (two sites with coverage under 100X in any sample, minimum coverage of any pathogenic variants site 826X in any sample) (**Fig. 5b, Supplementary** Fig. 9). Compared to the heteroplasmy levels elucidated from the day-3 biopsies by PCR-RFLP, the day-5 10X WGS-PGT results varied by 1.5% lower to 3.3% higher (**Fig. 5a**). Subsampling the sequencing data had little effect on the calculated heteroplasmy levels with estimates varying up to a maximum of 1.6% in any sample at different levels of genome-wide coverage (**Supplementary Table 6-7**). The subsampled 10X WGS-PGT heteroplasmy levels from the surplus embryo were 0% to 3.6% higher compared to the day-3 biopsy and between 1.6% lower and 1.5% higher compared to the day-5 biopsy (**Fig. 5a, Supplementary Table 6-7**). Similarly, when TE biopsy and surplus embryo were sequenced directly at 10X (*n* = 2, **Fig. 1b**), the heteroplasmy levels obtained for PCR-RFLP on day 3 differed by 4% from WGS TE biopsy result and there was a 1 to 1.8% difference between WGS results from the TE biopsy and the surplus embryo material (**Fig. 5a, Supplementary Table 8**). In two cases, where the day 3 biopsy with PCR-RFLP did not yield a result, a heteroplasmy level could be obtained using WGS-PGT.

**Fig. 5.**
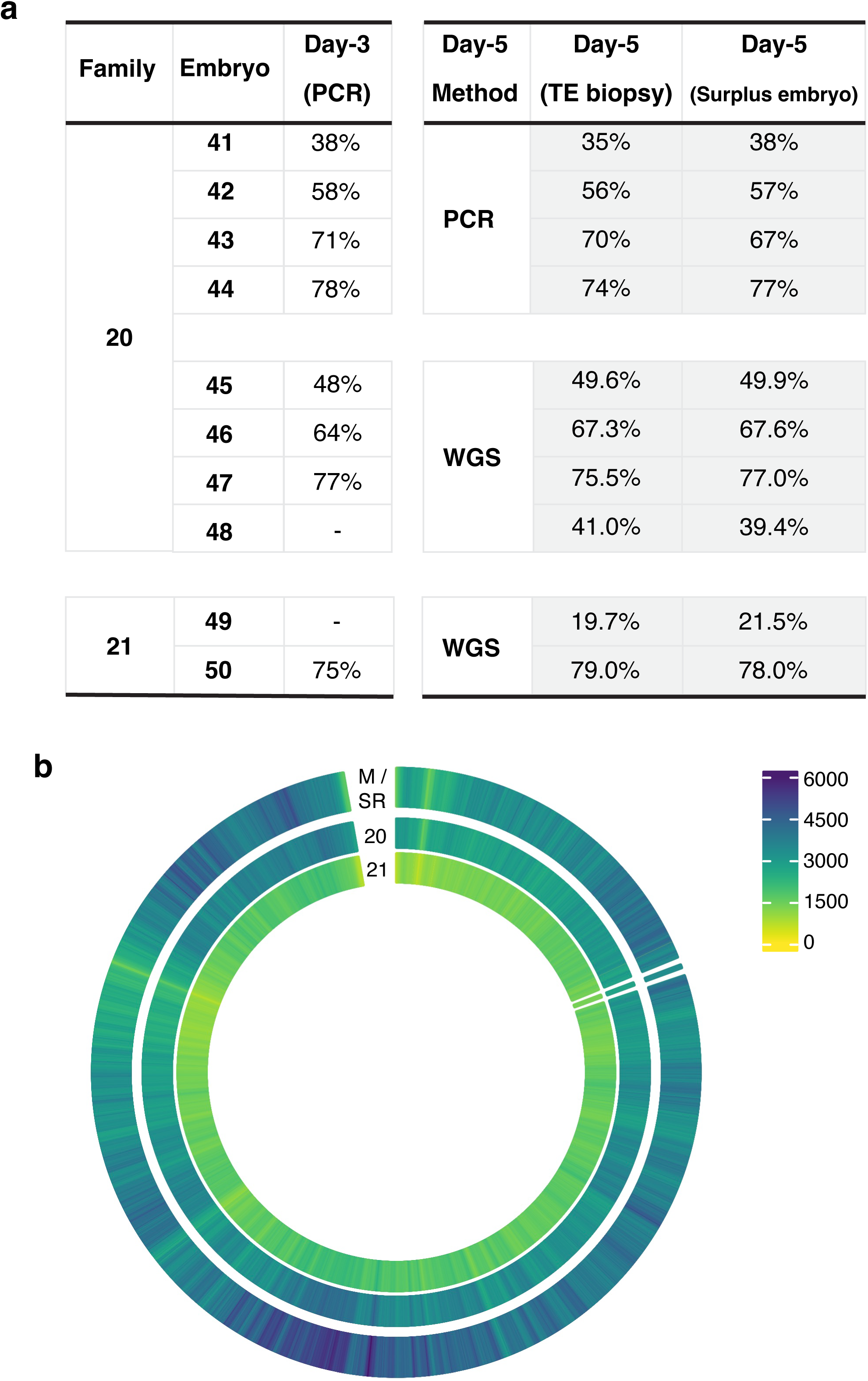
PGT for mtDNA disorders. **a,** Heteroplasmy levels in m.3243A>G embryos as determined by PCR-RFLP for day-3 blastomere biopsy, day-5 TE biopsy and surplus embryo material (embryos 41-44) or using ∼10X WGS-PGT (embryos 45-50). **b,** Mean depth of coverage per mtDNA position is shown for ∼10X WGS-PGT data from embryos sequenced for PGT-M or PGT-SR (outer ring, *n =* 23), deep sequencing with subsequent sub-sampling for PGT-MT (middle ring, *n* = 4) and direct ∼10X sequencing for PGT-MT (inner ring, *n =* 2). Position 0 is at the top with the positions ordered clockwise and colour representing the number of reads per position. The MELAS mutation position (m.3243) is highlighted as a separate segment and magnified 30x compared to all other positions. PGT: preimplantation genetic testing; MT: mitochondrial DNA disorders; RFLP: restriction fragment length polymorphism; TE: trophectoderm biopsy; WGS: whole genome sequencing; M: monogenic; SR: structural rearrangement; mtDNA: mitochondrial DNA.

## Discussion

We present WGS-PGT, a clinical whole genome sequencing method for all forms of PGT that outperforms traditional and state-of-the-art PGT technologies (**Fig. 6**). The advent of SNP-array and GBS-based PGT methodologies enabled generic assays, but these approaches still faced challenges in complex genomic regions, e.g. telomeres and centromeres, in consanguineous couples, and when there is haplotype recombination in proximity of the ROI. The increased resolution through WGS-PGT allowed more embryo samples to meet the diagnostic assessment criteria thresholds in these complex genomic regions for PGT-M. In parallel, PGT-M via haplotyping can be complemented with direct pathogenic variant detection. WGS-PGT enabled parents-only PGT-AO by identifying aneuploidies, their segregational and parental origin and level of mosaicism without the need of an available close-relative for phasing. We demonstrate the ability to detect structural rearrangements and distinguish normal from balanced embryos through both a direct approach, leveraging paired-end sequencing information from the embryo samples, and an indirect approach, which involves analyzing flanking haplotypes. Even though various strategies exist to differentiate normal from balanced embryos, including MaReCs^42^, a method based on shallow sequencing^43^, and a method based on Nanopore sequencing technology^44^, they cannot be integrated with other workflows for other PGT purposes as each workflow requires different technologies. Finally, we show the comparability and reproducibility of heteroplasmy levels between traditional day-3 blastomere biopsy with PCR-RFLP and our WGS-PGT method. We demonstrate that all mtDNA positions in the samples sequenced at 10X coverage have a mtDNA coverage of more than 100X except for one site. While prior studies have demonstrated the ability to identify mtDNA variants and heteroplasmy levels from WGS data^45,46^, integrating these into an all-in-one PGT method is an important step forward that makes PGT scalable and accessible.

**Fig. 6.**
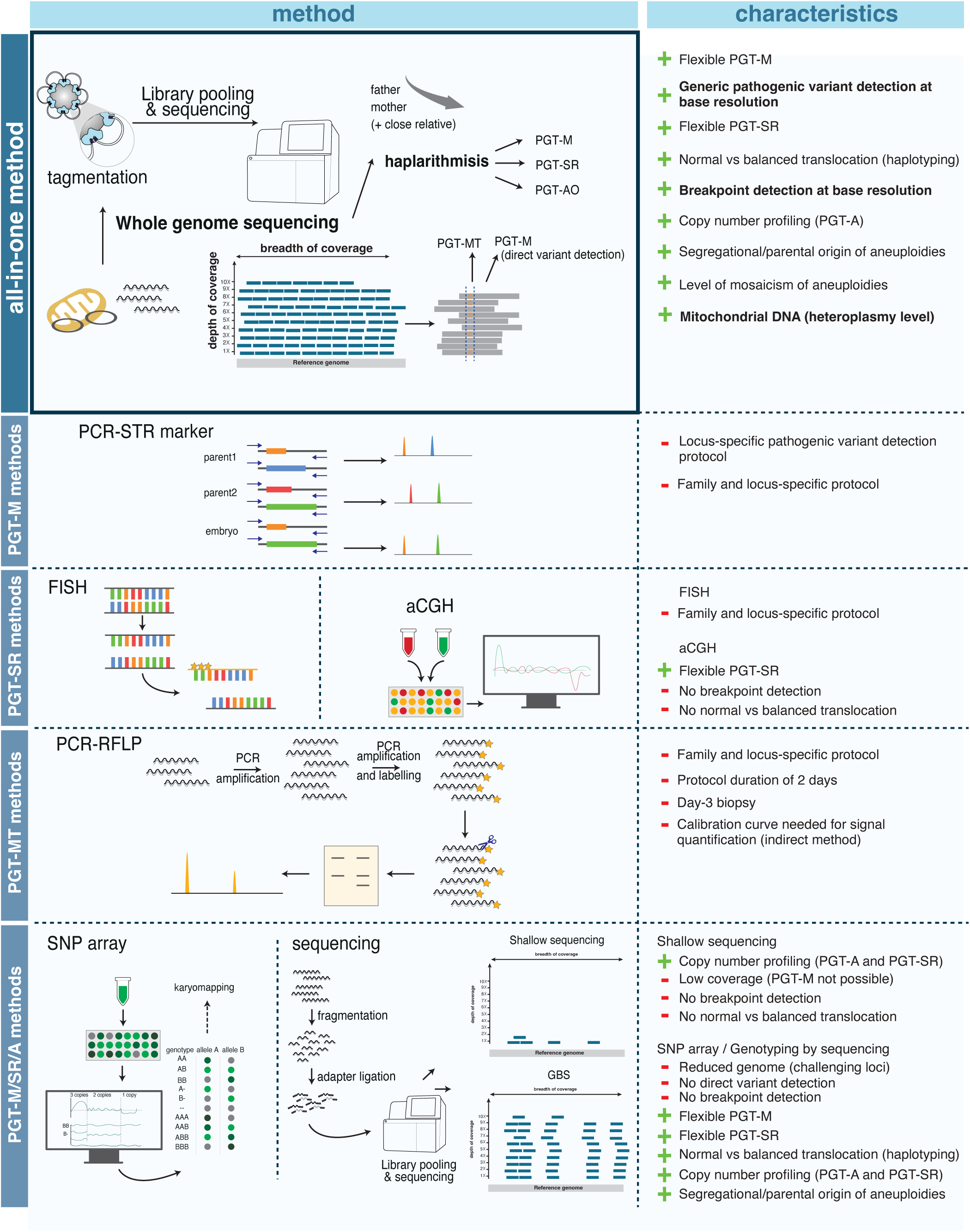
WGS-PGT as an all-in-one method. Illustrations and characteristics of PGT methods. WGS-PGT serves as an all-in-one method that captures all the functions within a single approach, while more traditional PGT methods are each designed for a specific purpose. WGS: whole genome sequencing; PGT: preimplantation genetic testing.

A key feature of WGS-PGT is the ability to directly detect the pathogenic variant of interest. Direct variant detection can complement haplarithmisis by resolving uncertain or inconclusive findings and by offering a solution in families with a *de novo* variant in one of the parents or when a suitable close relative for phasing is unavailable. In all cases where the depth of coverage exceeded 5X, we could correctly detect the SNV. A prior study that conducted WGS on embryos to identify *de novo* pathogenic variants for known diseases and polygenic risk score analysis^46^ did not account for allelic dropout or drop-in issues related to amplification as they did not include parental sequencing information in their analysis. An advantage of our method is that it can be performed both in a direct and indirect fashion using haplarithmisis principles, to be certain of our pathogenic variant of interest.

Apart from the primary objective of PGT-A – *i.e.* prioritising embryos with the highest implantation potential – we introduced PGT-AO, which determines the segregational origin of aneuploidies and their degree of mosaicism in parallel. The distinction between PGT-AO and PGT-A underscores the divergence between the objectives of selecting a chromosomally euploid embryo or an embryo with the highest implantation potential for transfer. Importantly, our approach permits the analysis of haploid or triploid embryos and embryos with meiotic aneuploidies, particularly those involving chromosomes assessed in non-invasive prenatal testing (NIPT)^47^. Mitotic aneuploid embryos are often mosaic and the abnormality may not be uniformly distributed throughout the blastocyst^48–51^. Furthermore, previous studies have shown that mosaic embryos can result in healthy offspring^26,27^, and chromosomal mosaicism may disappear via a self-correction mechanism where aneuploid cells are depleted in the inner cell mass and remain present in the TE lineage^52^. Current copy number quantitation methods fail to differentiate between true mosaic embryos and uniformly euploid or aneuploid embryos with technical noise^53^. These findings underscore the importance of assessing the parental and segregational origin of aneuploidies besides the degree of mosaicism. The ability of PGT-AO to select an embryo without meiotic aneuploidies, most likely helps couples from the difficult decision of potentially terminating a pregnancy of a foetus with a chromosomal aberration^54^. Longitudinal non-selection studies are needed to explore the connection between the segregational origin of aneuploidies implantation and viable pregnancy outcomes. This innovation raises ethical, legal, and social issues that need further reflection (**Supplementary Note ELSI**)

Our WGS-PGT method also have some limitations. The coverage levels may not suffice for accurate *de novo* pathogenic variants detection, which is recommended to be 30-40X^55^. To address this, we propose increasing depth of coverage to 30X when there is a *de novo* pathogenic variant in the parents. The genome-wide Mendelian inconsistency rate of 2.42%, representing both amplification errors, sequencing errors and putative *de novo* pathogenic variants. Notably, when specifically examining bulk samples from trios, others have observed Mendelian inconsistency rates of 1.92%^56^. These numbers highly exceed the average rate of *de novo* pathogenic variants of 1.20[]×[]10^−8^ per nucleotide per generation (∼0.3% per generation)^57^, probably attributable to sequencing errors. Longitudinal validation of the direct pathogenic variant detection is essential. Although incorporating PGT-MT within the same workflow is possible, we need further validation of WGS-PGT for mtDNA disorders on day-5 biopsies due to the limited sample size of PGT-MT families. Another limitation of our method and PGT in general is the need for embryo biopsy, as biopsy procedures demand specialised technical expertise, costly equipment and might impede embryo viability. In response, developments in the field of PGT have focused on utilising non-invasive DNA sources like cfDNA in the spent culture medium^58^ that originates from inner cell mass, TE cells^59^, cumulus cells, and polar bodies^60^. Potential contamination from cells or maternal origin should be assessed in future methods. Haplarithmisis can distinguish between contributions from maternal and fetal genomes in placental DNA samples^61^, therefore our WGS-PGT method could be used to tackle the maternal contamination and thus WGS-PGT is future-proof for development of niPGT.

In summary, we have presented a new strategy for all-in-one PGT that is able to capture all forms of PGT into one method. WGS-PGT enables a simplified, scalable, and universal PGT that outperforms current state-of-the-art PGT methods and has the capacity to enhance reproductive genetic care.

## Acknowledgements

We are grateful to all families that participated in this study. This study was funded by The EVA (Erfelijkheid Voortplanting & Aanleg) specialty program (grant no. KP111513), the Horizon Europe (NESTOR, grant no. 101120075) of the European Commission and the Horizon 2020 innovation (ERIN, grant no. EU952516) grants. We would like to specially thank F.J.M. Snepvangers, L.E.C. Meers, W. Loneus, K. Daenen, S. Spierts, B.M. Reuters and M. Kurvers-Alofs for their support with all clinical wet-lab related procedures. Also, we would like to thank R.C. Derks from the Radboudumc Genome Technology Center for WGS data generation.

## Methods

### Study participants and ethical approval

Couples were counseled by clinical geneticists at Maastricht University Medical Centre + (MUMC+) and enrolled in the diagnostic preimplantation genetic testing (PGT) procedure (licensed by the Dutch Ministry of Health, Welfare and Sport CZ-TSZ-291208) after signing an informed consent form. Couples who underwent PGT, consented for the use of affected embryos for the development of PGT methods. Full ethics committee approval was not required owing to the retrospective design of the study and the anonymized handling of the data under file number 2023-0091 from the ethics committee from the Maastricht UMC+. Genetic and clinical data shared in the context in this study cannot be used to identify individuals. We included 21 families who had undergone PGT for (double) monogenic, structural, or mitochondrial indications where spare (amplified) DNA from all samples was available. No additional biopsies were performed for this study specifically, except for PGT-MT (see in section PGT for mitochondrial DNA disorders).

### GBS-PGT sample collection and processing

For PGT-M the standard clinical procedure in our facility involved genotyping-by-sequencing (GBS), as described previously^62^. These procedures are part of standard clinical practice and were not performed for this study specifically. Briefly, peripheral blood samples were collected from prospective parents and close relatives from which DNA was isolated using the QIAsymphony DSP DNA Midi kit (Qiagen, Germany). A trophectoderm (TE) biopsy, *i.e.* 5-10 cells, was taken from sufficiently developed embryos on day-5 and the collected material was subjected to multiple displacement amplification (MDA) using the REPLI-g Single Cell kit (Qiagen, Germany), following the manufacturer’s instructions. Library preparation, using the OnePGT solution (Agilent Technologies) was then carried out on genomic DNA samples from parents and close relative(s), along with the whole genome amplified (WGAed) samples from embryos, following the manufacturer’s instructions and as previously described^62^. The libraries were sequenced on a NextSeq 500 sequencing system. All excess DNA not used for library preparation was stored at −20 ° C, in accordance with clinical standards.

### Whole genome sequencing sample processing

DNA from parents and close relative(s) and WGAed DNA from embryos, (see above) that was stored according to clinical standards, was subjected to whole genome sequencing (WGS) library preparation. Briefly, a minimum input of 20 μl, with a concentration of 30 ng/μl, was supplemented with 0.12 ng of embryo tracking system (ETS) fragments (concentration: 0.03 ng/μl)^63^. Subsequently, bead-linked transposome (BLT) PCR-free library preparation (Illumina, San Diego) was carried out according to the manufacturer’s instructions for input quantities ranging from 300 to 2000 ng. The resulting libraries were purified using a double-sided bead purification process. Sequencing was performed using a NovaSeq 6000 in the Radboudumc to a target depth of coverage of ∼30X-40X or 10X.

### Sequencing data processing and quality control

The raw sequencing data were demultiplexed and aligned to the human reference genome (complemented with the sequences of all ETS amplicons) using bwa-mem2 (v. 2.2.1)^64^. The WGS data were aligned to hg38 while the GBS data were aligned to hg37, and positions were then converted to hg38 using liftOver from the Rtracklayer package^65^. The quality of the resulting alignment was assessed using Qualimap (v.2.2.1.)^66^ to determine breadth and depth of coverage as well as the purity of the expected ETS fragment.

Samples that underwent deep (∼40X) sequencing, were subsampled using the “view” function from SAMtools (v. 1.15.1)^67^. The fraction of the original bam file required to generate different subsets was calculated by dividing the target coverage (5X, 10X, 20X and 30X) by the original coverage. Wilcoxon Rank-Sum Tests were used to compare the GBS and WGS groups and Kruskal-Wallis test with Dunn’s multiple comparisons test to compare the different target coverage groups.

### Haplarithmisis-based PGT

GBS and WGS data were analysed using a modified version of the siCHILD analytical pipeline that is equipped with haplarithmisis^68^ and has been further adapted for sequencing data^63^. Initially, a preparatory test was conducted for the parents and close relative(s) to assess whether the couple was eligible for PGT. Subsequently, an “embryo test” was run in which embryo haplotypes were reconstructed to ascertain a diagnosis. Using this pipeline, aligned sequencing data was processed using Joint HaplotypeCaller from GATK (v. 3.4-46)^69^ to extract the genomic locations in the dbSNP database (v. 150). All subsequent processing was carried out in R (version 3.3.1)^70^ as previously described^63,68^. Briefly, the GATK output was used to determine the genotype per position of each sample using vcfR (v. 1.8.0.9000)^71^ R package. This was used to calculate B allele frequencies (BAF), which were subsequently phased by leveraging information from the parents and a close relative. Segmentation of the parent specific phased BAFs was used to determine the haplotypes. Copy number states for 100kb-sized genomic bins were assessed using the QDNAseq (v. 1.10.0)^72^ R package and segmented using piecewise constant fitting (PCF)^73^ with a gamma value of 50. GBS-PGT requires eight informative SNPs at either 2Mb side of the region of interest (ROI) (**Supplementary Table 2**).

The proportion of these informative SNPs should be at least > 80% concordant with either the affected or unaffected haplotypes as determined by parental phasing.

### Haplarithmisis comparison between GBS and WGS

Haplarithmisis output was evaluated from subsampled data generated at ∼10X depth of coverage by assessing mendelian inconsistency level, number of informative SNPs, and haplotype concordance. The assessment criteria for the preparatory test and embryo test are listed (**Supplementary Table 2**). Mendelian inconsistency rate was defined as the proportion of inconsistent genotypes out of the total number of genotypes that were analysed. These rates were calculated for individual chromosomes and then the mean for all autosomes was calculated. Haplotype concordance between GBS and WGS was determined by comparing the interpreted haplotypes per parent. Subsequently, the haplotype concordance of WGS-PGT with GBS-PGT was assessed for each target coverage using Kruskal-Wallis test followed by Dunn’s multiple comparisons test.

### Direct mutation detection

Single nucleotide variants (SNVs) and deletions in affected embryos were evaluated by examining the nucleotides at the base-level. We analysed genetic loci that entailed single nucleotide alterations such as SNVs and deletions. In total 22 genetic loci were included, of which 19 SNVs and three deletions. Moreover, 11 genetic loci were analysed that entailed deletions ranging 2 bp from 398 kb. Reads mapping the relevant genetic location were extracted from bam files by indicating the chromosome, start and end position of a genomic interval using the SAMtools “view” function^67^. The resulting bam files were visualised in the Integrated Genomics Viewer (IGV) (version 2.11.9) to ascertain the nucleotides at the indicated position. PGT results determined by haplarithmisis (affected, not affected, carrier, inconclusive) were compared with the putative diagnosis based on direct variant detection. Moreover, the expected genotype based on the haplotyping result was compared to the genotype as ascertained by direct variant detection.

### PGT-AO: classification of (segmental) chromosomal abnormalities, their segregational origin and their level of mosaicism

Copy number variation (CNV) calls were visualised employing haplarithms. Copy number state of the embryos was determined by analysing log2 ratio of the observed copy number to the expected copy number, as indicated by logR ratios, alongside shifts in genotype frequencies of the reads, measured by BAFs. Subsequently, the aberrations were classified based on several criteria. (i) the copy number aberration detected (*i.e.* gain or loss), (ii) the size of the aberration (*i.e.* genome-wide, chromosomal, or sub-chromosomal), (iii) the parental origin of the aberration (*i.e.* paternal or maternal), (iv) the segregational origin of the aberration (*i.e.* meiosis I, meiosis II, or mitosis), and (v) the degree (>10%) of mosaicism. To determine the degree of mosaicism, the genomic coordinates at the logR shift were used to extract the segmented parental phased BAF of the location of interest. BAF values were then compared to the reference dataset by Conlin et al. 2010^74^. Besides conventional haplarithmisis that includes a close relative to phase the parents, “parents-only phasing” was performed by phasing the parental genome with the embryo itself.

### PGT for structural rearrangements

Three families with PGT-SR indications were included. Copy number variation was originally analysed using the Veriseq-PGS kit (Illumina Inc., Santa Clara, CA) according to the manufacturer’s instructions. For the family that only had a PGT-SR indication, an additional TE biopsy was taken from the affected surplus embryos to generate a WGAed DNA sample using the REPLI-g Single Cell kit (Qiagen, Germany). In the case of families with both PGT-SR and PGT-M indications, the excess WGAed DNA was used and re-processed with our WGS-PGT approach as described above. The data for families with dual PGT-M and PGT-SR indications (family 1 and 9), for which close relative(s) were also sequenced, were processed and visualised as described for PGT-M cases. Where deep sequencing (30-40X) was undertaken, the subsampling strategy was also applied, and structural rearrangements were assessed at all target coverages. For one family (family 19) no referent individual was sequenced, in this case each embryo was used to phase the remaining embryos. Derivative chromosome breakpoints were ascertained using Manta (v1.1.0) with default settings^75^. The resulting variants were then filtered to include only break points (“BND”) where pairs of “mates” were identified on the expected chromosomes. In cases where Manta did not identify identical breakpoints in the embryo, the breakpoints were estimated from the haplarithmisis output, specifically from the segmentation of the phased parental BAFs and the segmentation of the logRs.

### PGT for mtDNA disorders

10 embryos from 2 families were included (Fig. 1b) that were deemed affected (> transfer threshold 15%)^39,76^ for mitochondrial encephalopathy, lactic acidosis and stroke-like episodes (MELAS, m.3243A>G) based on results from the current gold standard blastomere-biopsy (day-3) testing. The embryos were re-biopsied on day-5 to obtain a TE biopsy sample and the remaining embryo defined as surplus embryo was also analysed to gain an accurate representation of the true heteroplasmy level. Four TE biopsies and their corresponding surplus embryos from family 20 were re-analysed using the PCR-based restriction fragment length polymorphism (PCR-RFLP) protocol that was also used to analyse the day-3 biopsy samples. The protocol was implemented as previously described by Sallevelt et al^39^. Briefly, the biopsy material was subjected to cell lysis followed by two rounds of PCR.

The first amplification PCR was carried out with unlabelled primers for the m.3243A>G mutation, after which a fluorescently labelled primer was added for the second PCR round. The resulting product was enzymatically digested, purified, and analysed by capillary electrophoresis. The mutation load was determined by dividing the area of the mutation peak by the sum of both the wild type and mutation peak. The remaining four embryos from family 20 (TE biopsy and surplus embryo) were processed with the WGS-PGT protocol described above. These samples underwent deep sequencing with a target sequencing depth of 30-40X. The sequencing data were processed and subsampled as described above. The two embryos from family 21 (TE biopsy and surplus embryo) were also processed with WGS-PGT and sequenced to a depth of ∼10X. Sequencing depth per position was determined using the “depth” function from SAMtools^67^. Known pathogenic variants in the mitochondrial genome were extracted from the MITOMAP’s confirmed pathogenic mutations database^77^. The “HaplotypeCaller” function form GATK was used to determine the number of reads supporting the reference and alternative alleles at the indicated position^78^, from which the heteroplasmy percentage was calculated.

### Data visualization

Data were visualised using R packages ggplot^79^, circlize^80^, ggpubr^81^, and cowplot^82^.

## Data Availability Statement

The whole genome sequencing data cannot be shared publicly to protect the privacy of the families that participated in the study. The anonymised data may be requested through the corresponding author and via application to data access committee of MUMC+. Processed data presented in this paper, is submitted as Supplementary Table 9.

## Code availability

The code of haplarithmisis and the scripts used in the analysis of this study are available via Github (https://github.com/CellularGenomicMedicine/WGSPGT).

