## Supplementary Information for "Clinical-grade whole genome sequencing-based haplarithmisis enables all forms of preimplantation genetic testing"

### Supplementary Material

**Supplementary Table 1 | Genetic Indications of families**

| Family | PGT Type | Indication | Inheritance Pattern | Carrier | Genetic Interval a | Genetic Interval b | Speciality |
| --- | --- | --- | --- | --- | --- | --- | --- |
| **Family 1** | PGT-M | TGM1 | AR | both | chr14:24259813-24259814 |  | Double indication (PGT-M + PGT-SR) |
|  | PGT-SR | t(8;16)(p21.3;q24) | NA | father |  |  |  |
| **Family 2** | PGT-M | MEFV | AR | both | chr16:3243407-3243408 | chr16:3243310-3243311 | Double indication (PGT-M + PGT-M), consanguineous |
|  | PGT-M | DMD | X-R | both | chrX:31657989-31836820 |  |  |
| **Family 3** | PGT-M | ARSA |  |  | chr22:50626228-50626229 |  | Consanguineous, telomere |
| **Family 4** | PGT-M | BRCA1 | AD |  | chr17:43094569-43094570 |  | Double indication (PGT-M + PGT-M) |
|  | PGT-M | OCA2 | AR | both | chr15:28081711-28081712 | chr15:27985101-27985102 |  |
| **Family 5** | PGT-M | FSHD | AD |  | chr4:189518883-189519279 |  | Telomeric and homology to region on chr10 |
| **Family 6** | PGT-M | D2HGDH | AR | both | chr2:241743754-241743755 |  | Consanguineous |
| **Family 7** | PGT-M | PLN | AD |  | chr6:118558961-118558963 |  | Double indication (PGT-M + PGT-M) |
|  | PGT-M | DMD | X-R | both | chrX:32644132-32645152 |  |  |
| **Family 8** | PGT-M | GNAS | AD |  | chr20:58910751-58910752 |  | Recombination in maternal haplotype |
| **Family 9** | PGT-M | BSCL2 | AD |  | chr11:62702493-62702494 |  | Double indication (PGT-M + PGT-SR) |
|  | PGT-SR | t(2;11)(q35;q23) | NA | mother |  |  |  |
| **Family 10** | PGT-M | BOR1 | AD |  | chr8:71215510-71215511 |  | Recombination in maternal haplotype in embryo |
| **Family 11** | PGT-M | SCN9A | AD |  | chr2:166195187-166375987 |  | Paternal mitotic trisomy in chromosome of interest for one selected embryo |
| **Family 12** | PGT-M | NF1 | AD |  | chr17:31261761-31261762 |  | Direct mutation possible? |
| **Family 13** | PGT-M | TMEM67 | AR | both | chr8:93786253-93786254 | chr8:93755039-93755044 | Recombination in paternal haplotype in embryo |
| **Family 14** | PGT-M | TSC2 | AD |  | chr16:2080365-2080366 |  | Meiotic trisomy in selected embryo |
| **Family 15** | PGT-M | HemA | X-R | both | chrX:155022473-155022474 |  | Telomeric |
| **Family 16** | PGT-M | GJB2 | X-R | both | chr13:20223037-20565037 | chr13:20189547-20189548 | Recombination in maternal and paternal haplotype |
| **Family 17** | PGT-M | SLS | AR | both | chr17:19543703-19751473 |  | Centromeric |
| **Family 18** | PGT-M | SHFM3 | AD |  | chr10:101227249-101624830 |  | Mitotic trisomy in selected embryo |
| **Family 19** | PGT-SR | t(3;13)(q27;q33) | - | father |  |  | Double indication (PGT-SR + PGT-SR), without close relative |
|  | PGT-SR | ins(10;7)  (p11.2;q32.1q36.2) | - | father |  |  |  |
| **Family 20** | PGT-MT | MELAS | - | mother | chrMT: m.3243 A > G |  | mitochondrial indication |
| **Family 21** | PGT-MT | MELAS | - | mother | chrMT: m.3243 A > G |  | mitochondrial indication |

**Supplementary Table 2 | Assessment criteria for key parameters**

The table lists the key parameters guidelines for GBS-PGT and WGS-PGT in diagnostic practice. Since the data generated with WGS has a higher resolution, the assessment criteria thresholds are more stringent than for GBS. The assessment criteria are subdivided for a preparatory analysis and an embryo analysis. A preparatory test is performed to predict whether the sequencing data from parents and close relative(s) have sufficient quality to make a diagnosis. If the preparatory test passes the threshold, the couple can proceed with PGT. The ROI is defined as 2Mb up- and downstream of the genomic location of the indication.

WGS: whole genome sequencing; GBS: genotyping-by-sequencing; PGT: preimplantation genetic testing; ROI: region of interest.

| Guideline | GBS-PGT | WGS-PGT |
| --- | --- | --- |
| **Preparatory analysis** |  |  |
| Total number of reads | ≥ 26.000.000* | ≥ 160.000.000* |
| Mendelian inconsistency rate |  | ≤ 2% |
| Informative SNPs | | |
| Number of genome-wide informative SNP positions | ≥ 45.000 | ≥ 600.000 |
| Density at ROI | ≥ 4/Mb | ≥ 40/Mb |
| **Embryo analysis** |  | |
| Total number of reads embryo |  | ≥ 90.000.000 |
| Mendelian inconsistency rate | ≤ 15% | ≤ 6% |
| Distance ROI to homologous recombination event | ≥ 1 Mb |  |
| Haplotyping criteria | | |
| Number and fraction concordant SNPs between ROI and homologous recombination | ≥ 10 AB SNP's ≥ 90% Conc. | ≥ 10 AB SNP's ≥ 90% Conc. |
| Concordant SNPs in the region of ROI ± 2 Mb | ≥ 80% | ≥ 80% |
| Concordant SNPs in the region of ROI + 2Mb | ≥ 70%, **and** ± 2 Mb ≥ 80% | ≥ 70%, **and** ± 2 Mb ≥ 80% |
| Concordant SNPs in the region of ROI – 2Mb | ≥ 70%, **and** ± 2 Mb ≥ 80% |  |
| Number of consecutive correct or discordant SNPs < 10 concordant SNPs away from the ROI | ≤ 5 SNPs and ≤ 100 kb | ≥ 70%, **and** ± 2 Mb ≥ 80% |

*if the number of reads in the preparatory preimplantation genetic test does not meet the specified assessment criteria thresholds, and is lower than the recommended number of 160,000,000 but the SNP density within ±2MB around ROI meets the criteria of ≥40/Mb, then the preparatory test can still be approved. Moreover, close-relatives should include both grandparents, the gene should not be located in a telomeric/centromeric region, and the gene should not be located in a “difficult-to-analyse” region such as homologous region or a region with pseudogenes.

**Supplementary Table 3: Number of informative SNPs at ROI**

Listing the number of informative SNPs 2 Mb up- and downstream the ROI for each genetic indication and the concordance with affected or unaffected maternal/paternal haplotypes.

**Sheet 1**) GBS-PGT, **sheet 2**) WGS-PGT.

WGS: whole genome sequencing; GBS: genotyping-by-sequencing; ROI: region of interest; SNP: single nucleotide polymorphism, PGT: preimplantation genetic testing.

##### Supplementary Table 4 | Chromosomal abnormalities with parental and segregational origin and degree of mosaicism (%) determined by haplarithmisis

| Family | Embryo | Aberration* | chr | Parental origin | Segregational origin | Degree of mosaicism |
| --- | --- | --- | --- | --- | --- | --- |
| **Family 1** | 1 | Trisomy | 2 | Paternal | Mitosis | 30% |
|  | 4 | Monosomy | 19 | Maternal | ND | 100% |
| **Family 4** | 11 | Monosomy | 18 | Maternal | ND | 45% |
| **Family 7** | 18 | Trisomy | 10 | Maternal | ND | 25% |
| **Family 8** | 22 | Trisomy | 16 | Maternal | Meiosis I | 90% |
|  |  | Trisomy | 21 | Maternal | Meiosis II | 100% |
| **Family 9** | 25 | Monosomy | 2 | Paternal | ND | 100% |
|  |  | Trisomy | 11 | Maternal | Meiosis II | 100% |
| **Family 11** | 28 | Trisomy | 2 | Paternal | Mitosis | 100% |
|  |  | Monosomy | 8 | Maternal | ND | 100% |
|  |  | Trisomy | 14 | Paternal | Meiosis I | 100% |
| **Family 12** | 30 | Trisomy | 16 | Maternal | Meiosis I | 100% |
| **Family 14** | 32 | Trisomy | 4 | Maternal | Meiosis I | 80% |
|  |  | Trisomy | 16 | Maternal | Meiosis I | 100% |
| **Family 17** | 35 | Monosomy | 19 | Paternal | ND | 100% |
| **Family 18** | 37 | Triploidy | All  (except 15) | Maternal | Meiosis II | 100% |
|  |  | Disomy/trisomy | 15 | - | - | 50% |

##### Supplementary Table 5 | PGT-SR extended information

Tables with breakpoint pairs identified by Manta in the carrier parent and embryos of families with translocations. Karyotyping results, a schematic of the translocation and breakpoints estimated from VeriSeq PGT-SR are also included for each family. **Sheet 1**) Family 1, **sheet 2**) family 19, **sheet 3**) family 9, **sheet 4**) Copy Number Plots. From top to bottom, paternal haplarithms, maternal haplarithms, logR profile from QDNAseq and copy number profiles obtained from VeriSeq. In the paternal haplarithm and maternal haplarithm, each point indicates the parentally informative SNPs in blue (M1/P1) and red (P2/M2). The segmented parentally informative SNPs are shown as darkblue (M1/P1) or darkred (M2/P2). In the copy number profile from QDNAseq each point indicates the logR value (log2 copy number ratio between observed and expected) with a bin size of 100 kb and the segmented logR is depicted in orange. In the VeriSeq track, each point represents the copy number value of a bin sized ~1Mb. The green and red lines indicate the threshold for detection of copy number gain or loss, respectively.

PGT: preimplantation genetic testing: SR: structural rearrangements; SNP: single nucleotide polymorphism.

##### Supplementary Table 6 | PGT-MT number of reads at the MELAS (m.3243A>G) indication at different depths of coverage

Numbers shown represent the total number of reads at the site and the number of reads with the mutation in brackets. Embryos 45-48 from family 20 are shown.

PGT: preimplantation genetic testing; MT: mitochondrial DNA disorders; TE: trophectoderm; MELAS: mitochondrial encephalomyopathy, lactic acidosis and stroke-like episodes.

| Depth | Embryo 45 | |  | Embryo 46 | |  | Embryo 47 | |  | Embryo 48 | |
| --- | --- | --- | --- | --- | --- | --- | --- | --- | --- | --- | --- |
|  | TE  biopsy | Surplus  embryo |  | TE  biopsy | Surplus  embryo |  | TE  biopsy | Surplus  embryo |  | TE  biopsy | Surplus  embryo |
| Full | 11123 | 17205 |  | 11559 | 9287 |  | 8160 | 6952 |  | 6789 | 11559 |
|  | (5521) | (8517) |  | (6411) | (6287) |  | (6169) | (5303) |  | (2773) | (4464) |
| 30X | 4210 | 4634 |  | 4176 | 3996 |  | 3920 | 6952 |  | 3656 | 4333 |
|  | (2106) | (2283) |  | (2833) | (2743) |  | (2955) | (5303) |  | (1467) | (1742) |
| 20X | 3602 | 4191 |  | 3575 | 3346 |  | 3180 | 2956 |  | 2952 | 3776 |
|  | (1792) | (2076) |  | (2383) | (2301) |  | (2399) | (2228) |  | (1194) | (1473) |
| 10X | 2373 | 2965 |  | 2415 | 2231 |  | 2134 | 1972 |  | 1879 | 2472 |
|  | (1178) | (1481) |  | (1626) | (1509) |  | (1612) | (1518) |  | (771) | (974) |
| 5X | 1366 | 1831 |  | 1478 | 1288 |  | 1250 | 1135 |  | 1058 | 1488 |
|  | (675) | (910) |  | (977) | (885) |  | (936) | (872) |  | (436) | (586) |

##### Supplementary Table 7 | PGT-MT heteroplasmy levels for the MELAS (m.3243A>G) mutation at different depths of coverage

Embryos 45-48 from family 20 are shown.

PGT: preimplantation genetic testing; MT: mitochondrial DNA disorders; TE: trophectoderm; MELAS: mitochondrial encephalomyopathy, lactic acidosis and stroke-like episodes.

| Depth | Embryo 45 | |  | Embryo 46 | |  | Embryo 47 | |  | Embryo 48 | |
| --- | --- | --- | --- | --- | --- | --- | --- | --- | --- | --- | --- |
|  | TE  biopsy | Surplus  embryo |  | TE  biopsy | Surplus  embryo |  | TE  biopsy | Surplus  embryo |  | TE  biopsy | Surplus  embryo |
| Full | 49.6% | 49.5% |  | 67.1% | 67.7% |  | 75.6% | 76.3% |  | 40.8% | 38.6% |
| 30X | 50.0% | 49.3% |  | 67.8% | 68.6% |  | 75.4% | 76.3% |  | 40.2% | 40.2% |
| 20X | 49.8% | 49.6% |  | 66.7% | 68.85 |  | 75.4% | 75.6% |  | 40.4% | 39.0% |
| 10X | 49.6% | 49.9% |  | 67.3% | 67.6% |  | 75.5% | 77.0% |  | 41.0% | 39.4% |
| 5X | 49.5% | 49.7% |  | 66.1% | 68.7% |  | 74.9% | 76.9% |  | 41.2% | 39.4% |

##### Supplementary table 8 | Extended PGT-MT results for family 21 - mutation: MELAS (m.3243A>G)

TE PCR = RFLP-PCR applied directly to TE biopsy material, TE MDA-PCR = RFLP-PCR applied to WGAed TE biopsy material. WGS coverage shows number of reads at the indication site and number of reads with the mutation in brackets.

PGT: preimplantation genetic testing; MT: mitochondrial DNA disorders; TE: trophectoderm; MELAS: mitochondrial encephalomyopathy, lactic acidosis and stroke-like episodes; WGS: whole genome sequencing; MDA: multiple displacement amplification; WGAed: whole genome amplified; RFLP: restriction fragment length polymorphism; PCR: polymerase chain reaction.

| **Embryo** | **Sample type** | **TE PCR** | **TE WGA-PCR** | **WGS coverage** |
| --- | --- | --- | --- | --- |
| **49** | TE biopsy | 23% | 21% | 1424 (281) |
|  | Surplus  embryo | - | - | 5473 (1174) |
| **50** | TE biopsy | 77% | 76% | 1304 (777) |
|  | Surplus  embryo | - | - | 985 (1017) |

**Supplementary Table 9 | Assessment key parameters**

Tables with key parameters. **Sheet 1**) QC of PGT-M and PGT-SR samples at 10X coverage. **Sheet 2**) QC of sub-sampled samples from family 1 and family 2. **Sheet 3**) QC of PGT-MT samples.

PGT: preimplantation genetic testing; M: monogenic; SR: structural rearrangements; MT: mitochondrial DNA disorders; QC: quality control.

**Supplementary Figure 1 | Depth of coverage in the pilot study**
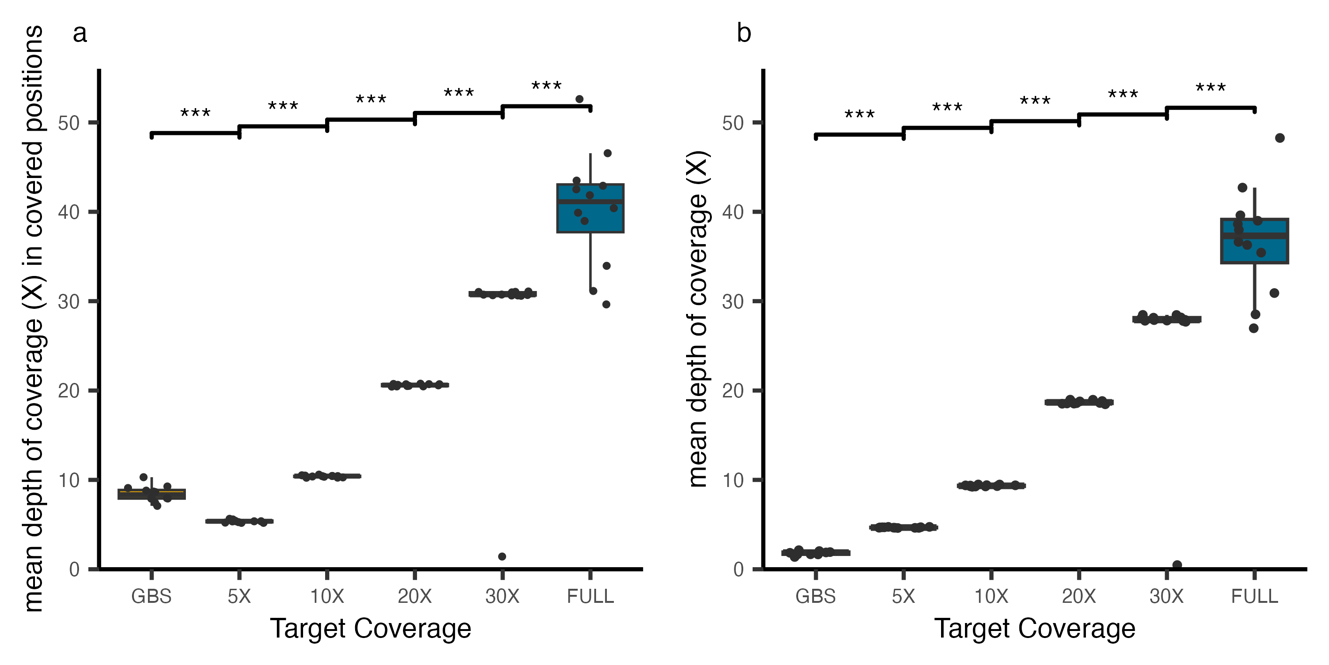

##### Depth of coverage for bulk DNA and WGAed embryo samples whole genome sequenced at an original sequencing depth (FULL: 30-40X) and corresponding *in silico* subsampled sequencing data at target coverages of 5X, 10X, 20X, 30X and GBS samples. The target depths of coverage (x-axis), and corresponding mean depth of coverage per sample (y-axis) are shown. a, The mean depth of coverage is calculated by dividing the number of bases of aligned reads by genomic positions that are covered by at least one read. b, the mean depth of coverage is calculated by dividing the number of bases of aligned reads by all genomic positions. The horizontal lines of the boxplots represent the 25^th^ percentile, median and 75^th^ percentile and the whiskers extend to 1.5 * IQR. The dots represent individual samples and WGS-PGT and GBS-PGT data is shown in blue and gold, respectively.

WGS: whole genome sequencing; GBS: genotyping-by-sequencing; IQR: interquartile range; PGT: preimplantation genetic testing.

##### Supplementary Figure 2 | Mendelian inconsistency

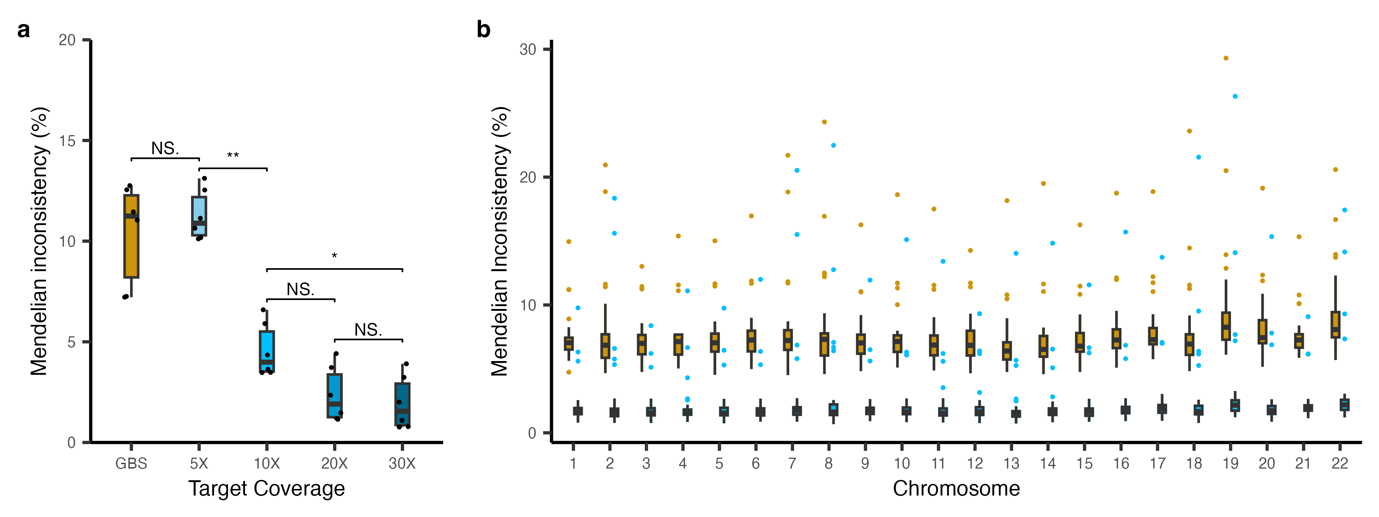

Mendelian inconsistency rates represent the proportion of genotypes in the embryo that violate Mendelian inheritance patterns from the total number of genotypes analysed. **a,** Mendelian inconsistency rates for the autosomes from *in silico* subsampled WGS-PGT data and corresponding GBS-PGT data (*n* = 6). The target depths of coverage (x-axis), and Mendelian inconsistency rates (y-axis) are shown. **b,** Mendelian inconsistency rates from WGS-PGT data sequenced at 10X and corresponding GBS data (*n* = 31) shown per autosomal chromosome (chr1-22). The horizontal lines of the boxplots represent the 25^th^ percentile, median and 75^th^ percentile and the whiskers extend to 1.5 * IQR. The dots represent individual samples and GBS-PGT and WGS-PGT data is shown in gold and blue, respectively.

WGS: whole genome sequencing; PGT: preimplantation genetic testing; GBS: genotyping-by-sequencing; IQR: interquartile range.

##### Supplementary Figure 3 | Haplotype concordance

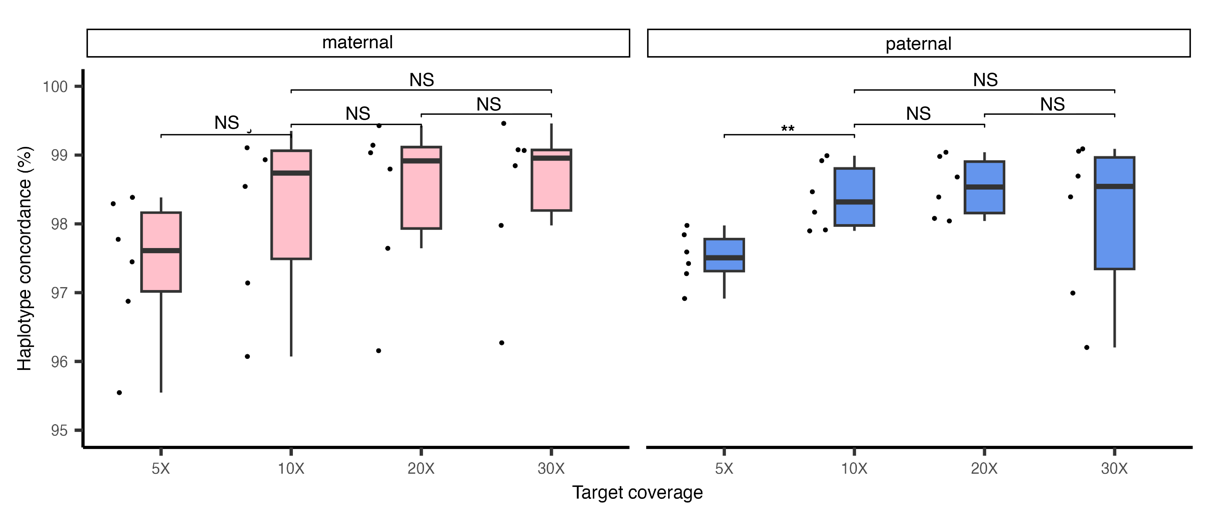

Haplotype concordance between WGS-PGT data and GBS-PGT data at different *in silico* subsampled depth of coverages (*n* = 6) for maternal (pink) and paternal (blue) haplotypes. The horizontal lines of the boxplots represent the 25^th^ percentile, median and 75^th^ percentile and the whiskers extend to 1.5 * IQR. The dots represent individual samples and maternal and paternal haplotype concordance is shown in pink and blue, respectively.

WGS: whole genome sequencing; PGT: preimplantation genetic testing; GBS: genotyping-by-sequencing; IQR: interquartile range.

**Supplementary Figure 4 | Informative SNPs**

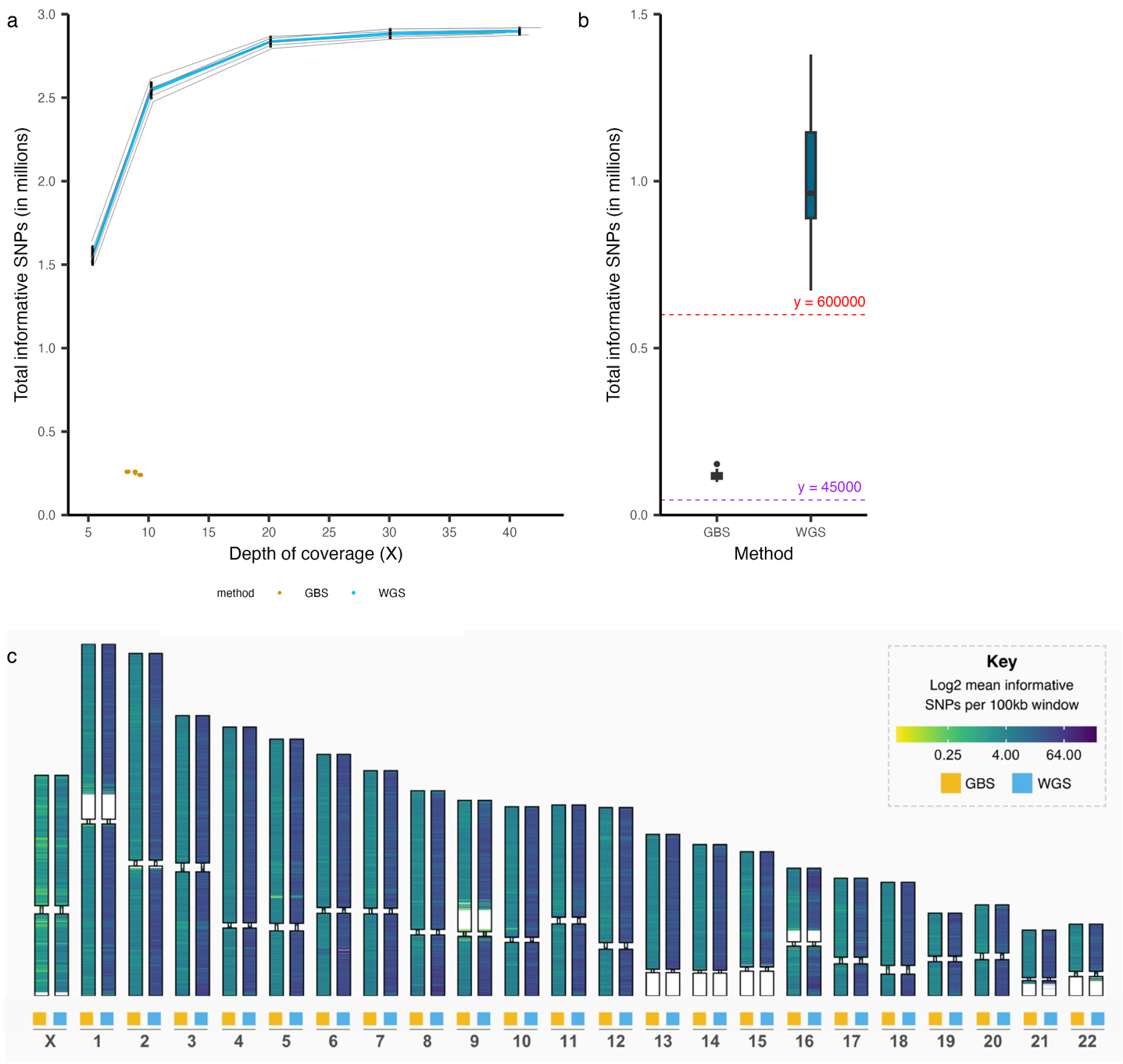

**a,** Total number of informative SNPs per embryo (*n* = 6) at different *in silico* subsampled depth of coverage from WGS data (lines: blue) and the corresponding GBS data (dots: gold). **b,** Total number of informative SNPs for GBS-PGT (gold) and WGS-PGT (blue). The dashed horizontal lines indicate the minimum required number of informative SNPs according to the quality control criteria for GBS (45.000 informative SNPs, purple) and WGS (600.000 informative SNPs, red). The horizontal lines of the boxplots represent the 25^th^ percentile, median and 75^th^ percentile and the whiskers extend to 1.5 * IQR. **c,** Heatmap showing the mean number of informative SNPs per phased parental haplotype in 100kb bins across chromosomes in data obtained by GBS or WGS. The mean number of informative SNPs was calculated from 51 phased parental (maternal or paternal) haplotypes from 31 embryos.

SNP: single nucleotide polymorphism; WGS: whole genome sequencing; PGT: preimplantation genetic testing; GBS: genotyping-by-sequencing; IQR: interquartile range.

##### Supplementary Figure 5 | Number of informative SNPs at ROI

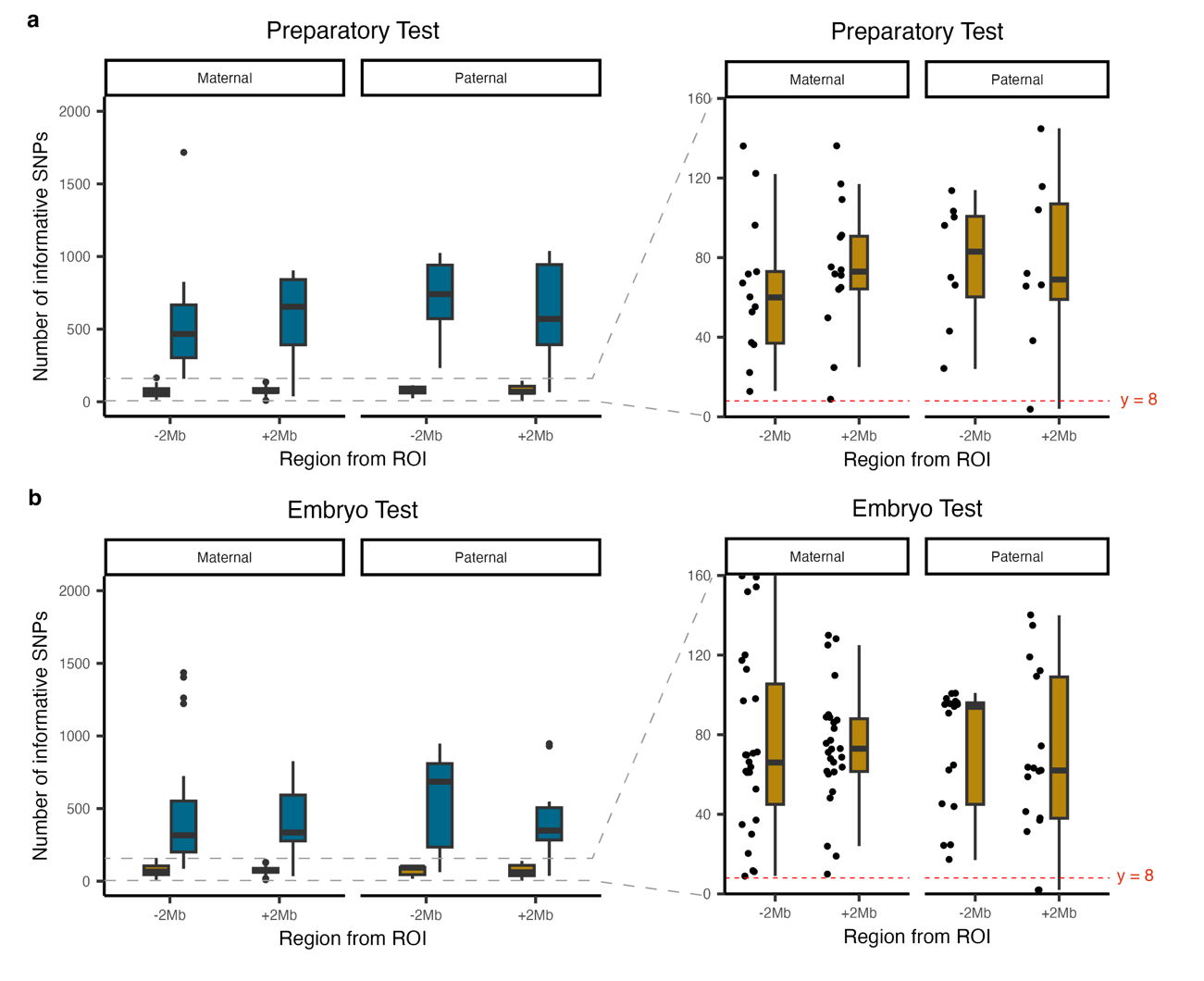

The number of informative SNPs 2 Mb up- and downstream of the ROI for WGS data (blue) and GBS (gold) data. Informative SNPs are categorised into maternal and paternal categories. Right panels: zoom-in on the number of informative SNPs for GBS-PGT data, the red dashed horizontal line represents the assessment criteria threshold for the minimum number of informative SNPs required for the region 2 Mb up- and downstream of the ROI. The horizontal lines of the boxplots represent the 25^th^ percentile, median and 75^th^ percentile and the whiskers extend to 1.5 * IQR. Dots represent number of informative SNPs for a single embryo. **a,** presents the number of informative SNPs for the preparatory test. **b,** presents the number of informative SNPs for the embryo test.

SNP: single nucleotide polymorphism; WGS: whole genome sequencing; GBS: genotyping-by-sequencing; ROI: region of interest; IQR: interquartile range; PGT: preimplantation genetic testing.

##### Supplementary Figure 6 | IGV sessions for single nucleotide direct mutation visualization

Each panel provides a comprehensive view of the genetic locus per family displaying single base pair substitutions and deletions. The family number and embryo numbers are indicated on the left side. At the top of each panel, an ideogram of the relevant chromosome is depicted. The precise location of the analysed window on chromosome scale is marked in red. A horizontal span arrow illustrates the width of this window, with the specific number of base pairs indicated. Within each panel, two tracks are featured for every embryo: the upper track serves as a coverage track, the lower track displays the reads. Nucleotides that match the reference genome are depicted as grey reads. Positions that do not match the reference genome are color-coded (A: green, C: blue, G: orange, T: red or deletion: short black horizontal line). The reference genome track, utilising the hg38 reference genome is positioned at the bottom for reference. Shown is WGS-PGT data at ~10X coverage.

IGV: integrative genomics viewer; WGS: whole genome sequencing; PGT: preimplantation genetic testing.

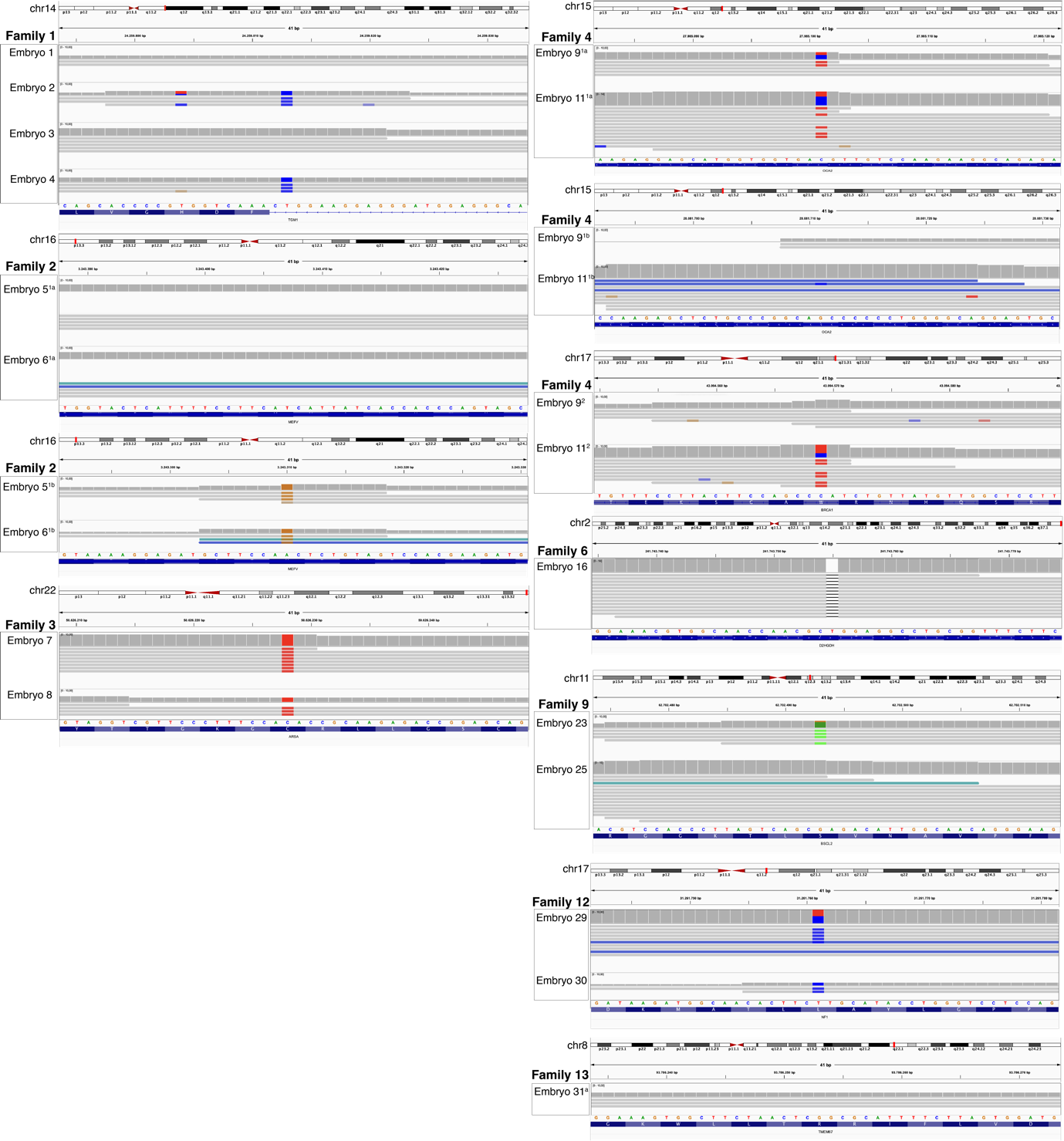

**Supplementary Figure 7 | IGV sessions for direct deletion visualization**

Each panel provides a comprehensive view of the genetic locus per family displaying deletions greater than 1 base pair. The family number and embryo numbers are indicated on the left side. Moreover, the haplarithmisis diagnostic result that belongs to this ROI of the embryo is presented along. At the top of each panel, an ideogram of the relevant chromosome is depicted. The precise location of the analysed window on chromosome scale is marked in red. A horizontal span arrow illustrates the width of this window, with the specific number of base pairs indicated. Within each panel, two tracks are featured for every embryo; the upper track serves as the coverage track, the lower track displays the reads. **a,** panels for embryos from families with a deletion ranging 2-3 base pairs. Deletions are visualised as short black horizontal line. **b,** panels for embryos from families with an autosomal recessive deletion. Deletions are visualised as loss of coverage. **c,** panels for embryos from families with autosomal dominant or x-linked deletions. Deletions are visualised using option view as pairs, where the coloured horizontal line connects the read pairs and is spanning the size of the deletion. The dashed red line indicates the size of the (undetected) putative deletion in embryo 5 from family 2.

IGV: Integrated genomics viewer; ROI: region of interest.

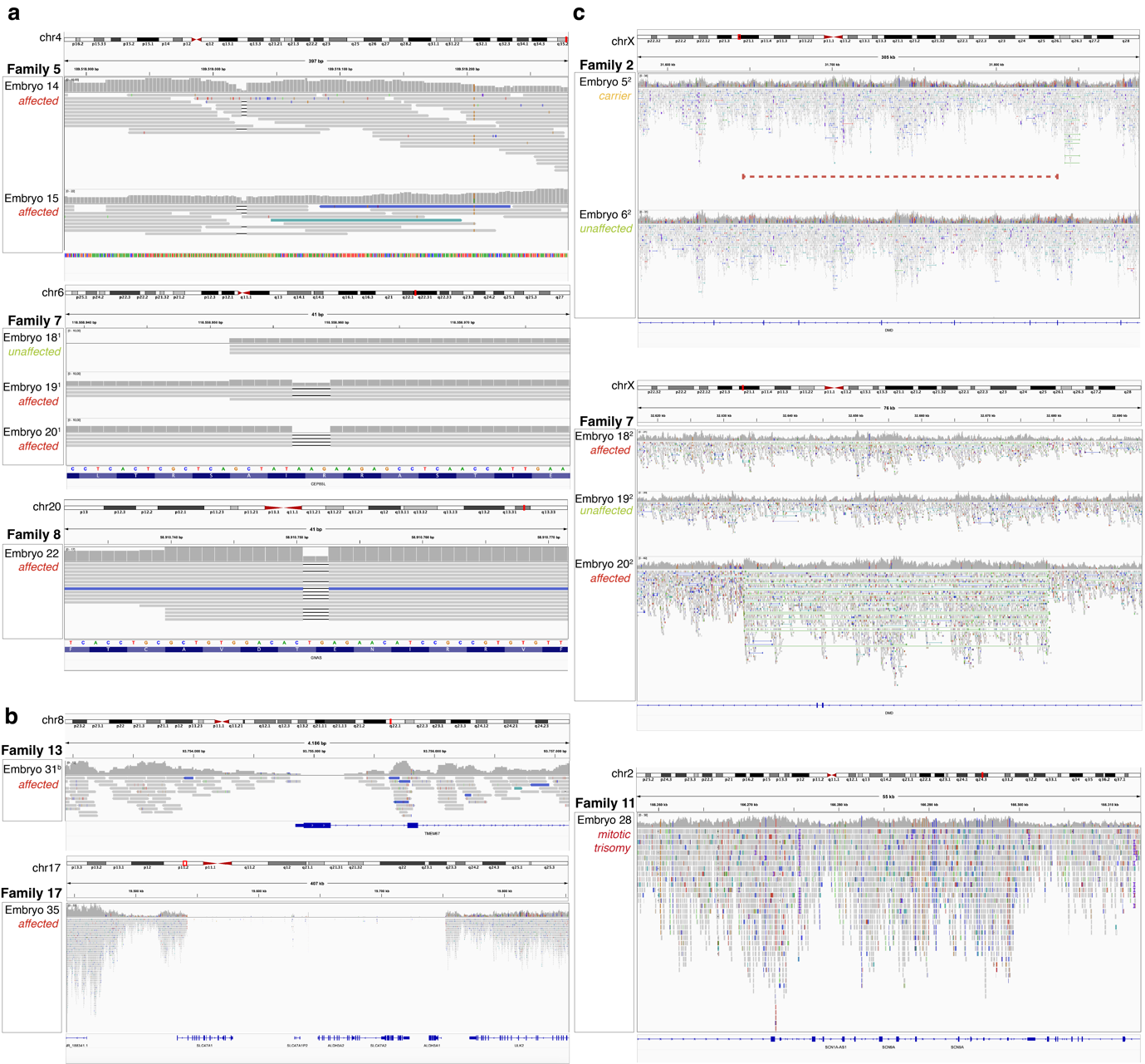

**Supplementary note Figure 3a: Parents-only haplarithmisis for PGT-AO**

Haplarithmisis is a conceptual workflow that enables simultaneous genome wide haplotyping and copy number typing using genotyping information from offspring (embryos), parents and a close-relative such as grandparents or a sibling^1^. To validate parents-only haplotyping and its determination of segregational origin, (i) the genotype of close-relatives was used as a seed for phasing and the segregational origin of copy number gains was determined, (ii) the genotype of the embryo itself was used as a seed for phasing and segregational origin of the same aberrations was determined (Fig. 3a).

Using conventional close-relative phasing, the segregational origin of aberrations can be determined by interpreting the parity and reciprocity of features for detection of segregational origin as described previously in detail^1^ (Fig. 3a). When a copy number increase involves a combination of different homologous chromosomes of one parent, this constitutes a meiotic error. If the centromeric regions originate from different homologues (e.g. maternal H1 and maternal H2), this indicates a meiotic I error. Alternatively, if the centromeric regions indicate the same homologue (e.g. maternal H1 and maternal H1), this indicates a meiotic II error. In contrast to meiotic errors, mitotic errors consist of an exact duplication of a single homologue. Distinguishing between meiotic II and mitotic error is only possible when a crossover event has occurred in the aberrant chromosome. A paternal trisomy involves two paternal and one maternal chromosome, while a maternal trisomy has two maternal and one paternal chromosome. Conversely, a paternal monosomy retains the paternal chromosome and expels the maternal one, and a maternal monosomy retains the maternal chromosome while expelling the paternal one.

For parents-only haplotyping, the same ruleset applies. However, the phasing is now limited to only include parental inheritance (paternal or maternal), instead of specific parental homologue inheritance (Paternal H1 or H2, maternal H1 or H2). Therefore, haplarithmisis categorises the parental chromosomal constitution (i.e., 1_paternal_:2_maternal_ allelic ratio) but cannot further categorise P1, P2, M1, and M2 B-allele frequencies (BAFs) as the specific parental homologue information is missing. Thus, in case of a maternal meiotic trisomy the segmented M1 BAFs (blue lines maternal haplarithm) will remain at a value of 0 instead of a value of 0.33 as compared to close-relative phasing. The segmented M2 BAFs (red lines maternal haplarithm) are consistent with segmented M2 BAFs of close-relative phasing. In case of a maternal mitotic trisomy, a crossover event is not recognised in parents-only phasing, placing the segmented M1 and M2 BAFs at 0 and 0.33 respectively. Importantly, the inconsistencies of segmented M1 and P1 BAFs between parents-only and close-relative phasing do not hamper determination of segregational origin, since segmented M2 and P2 BAFs are sufficient for this purpose.

##### Supplementary Figure 8 | PGT-SR segmental deletion and duplication detection at different depths of coverage

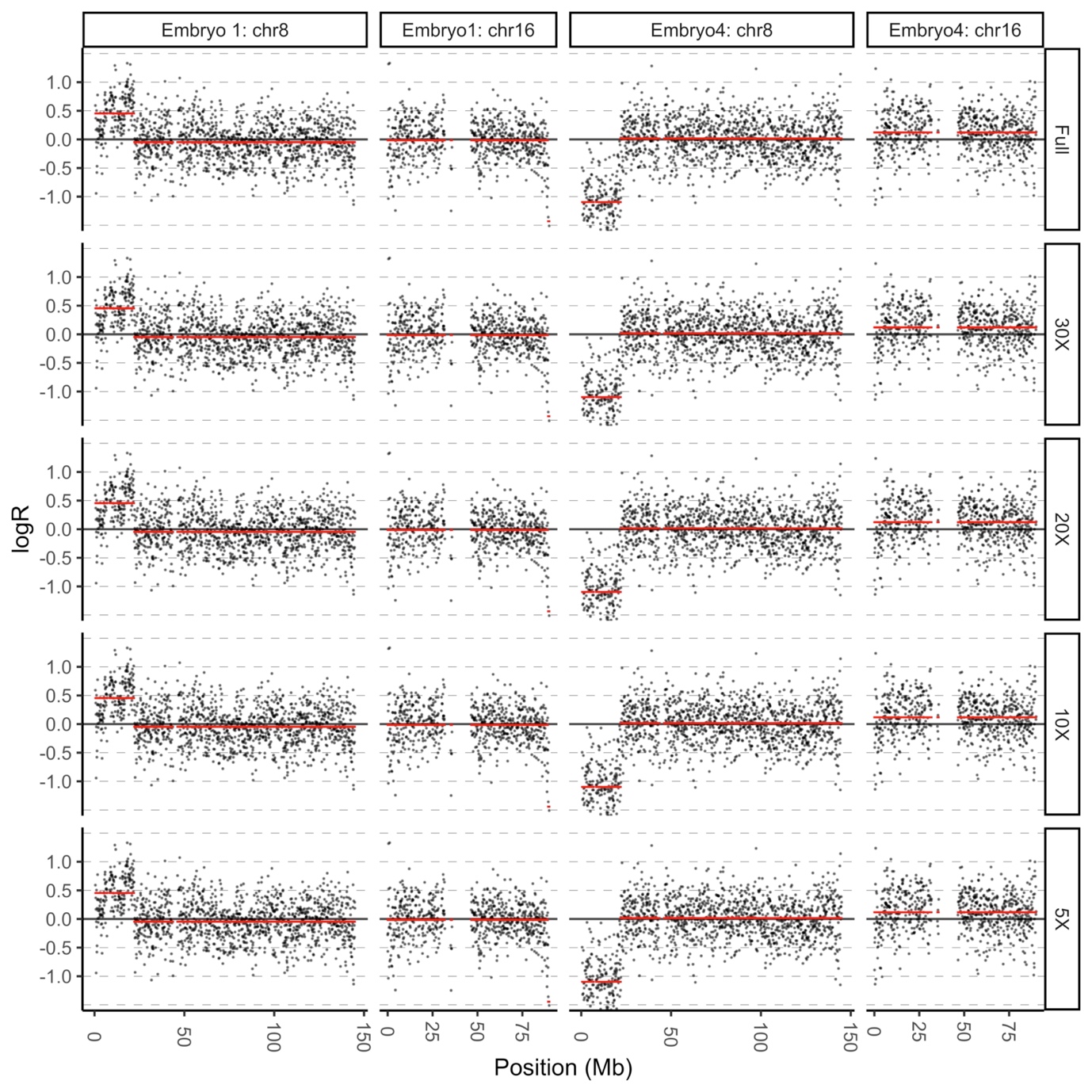

Raw logR values (black dots) and segmented logR values (red) derived from *in silico* subsampled sequencing data obtained from the original WGS-PGT data (FULL; ~30-40X). Two representative embryos (embryos 1 and 4) from a family with a paternal translocation of chromosomes 8 and 16 (family 1) are shown.

PGT: preimplantation genetic testing; SR: structural rearrangement; WGS: whole genome sequencing.

##### Supplementary Figure 9 | PGT-MT depth of coverage at mitochondrial pathological mutation sites at different depths of coverage

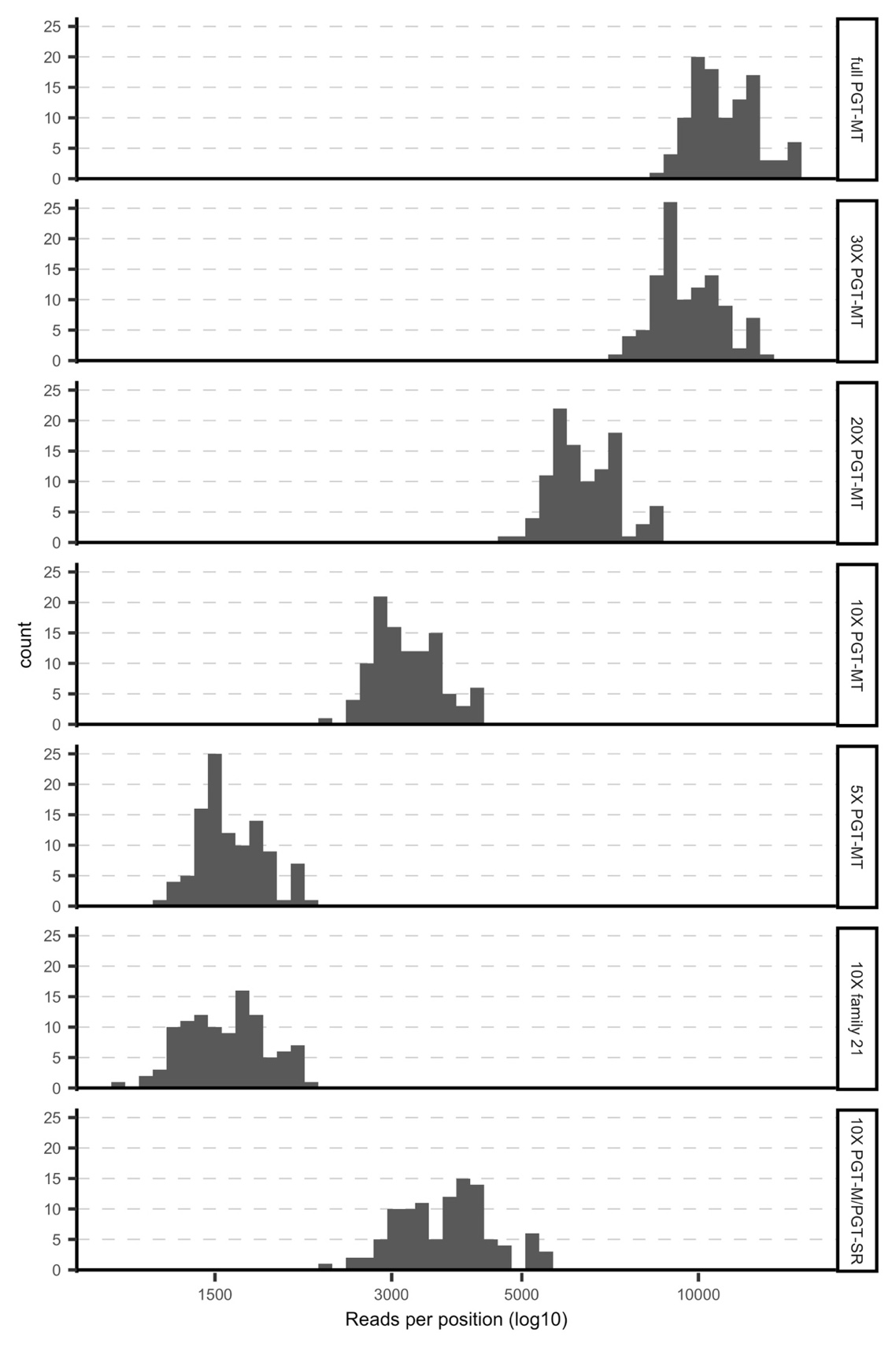

Histogram showing the mean number of reads per position of pathogenic mtDNA variants (*n* = 105) at different *in silico* subsampled depths of coverage as indicated (right side labels). The mean is calculated from four PGT-MT TE-biopsy samples (rows 1-5), two PGT-MT TE-biopsy samples (for 6) or 23 embryos sequenced at 10X for PGT-M / PGT-SR indications (row 7). Data from WGS-PGT is shown.

PGT: preimplantation genetic testing; MT: mitochondrial DNA disorders; TE: trophectoderm.

**Supplementary Note, ELSI section**

**Ethical, legal, and social implications**

Though the overall performance of WGS-PGT is promising, this innovation raises ethical, legal, and social issues (ELSI) that need further reflection. As additional testing for e.g. aneuploidy (PGT-AO) or *de novo* mutations entails screening, the three ethical screening criteria apply (1):

First, proportionality, i.e., the requirement that the screening’s possible advantages for the members of the target group should clearly outweigh its possible disadvantages. While possible benefits of PGT-AO in terms of preventing implantation failure, miscarriage or an affected ongoing pregnancy and related challenging decision-making may be substantial, there is no strong evidence yet regarding its clinical utility and proportionality. Clearly, PGT-AO introduces a layer of complexity when confronting mitotic-origin aneuploidy that is often mosaic. The decision of whether to undergo embryo ranking adds to the emotional weight of an already challenging reproduction journey. The offer of such screening, should, therefore, be embedded in further (pre-clinical and clinical), multi-disciplinary research. This cautious route is even more important regarding whole genome sequencing and analysis (WGSA) aimed at screening for *de novo* mutations, given the additional ethical and psychological complexities linked with e.g. (trio-)testing also for late-onset disorders. Considering the possibility of implementing embryo ranking, the absence of well-defined guidelines will place experts in a challenging decision-making role, one that can significantly influence family planning of couples and their overall well-being. Furthermore, the inclusion of additional criteria beyond the initial referral indications may increase the likelihood of discarding potentially healthy embryos, leaving prospective parents with the distressing situation of having a limited number of transferable embryos for IVF consideration.

Second, respect for patients’ (reproductive) autonomy. This requires (valid) consent, based on adequate information about the aims and present challenges and limitations of different applications of WGS-PGT. Regarding PGT-AO, information about the center’s policy regarding both reporting (different grades of) mosaicism and the (non-)transfer and ranking of embryos with putative aneuploidy of either meiotic or mitotic origin is crucial. The center’s handling of possible tensions between respect for autonomy on the one hand and the responsibility of doctors involved in reproductive medicine to also take account of the welfare of the possible future child thus conceived on the other must be addressed in counseling. Clearly, this issue is relevant for PGT more generally.

Third, justice, a criterion that is especially relevant for the possible implementation of WGS-PGT in collectively funded health care systems. Given the possible scarcity of financial resources for health care, may WGS-PGT be among the priorities to be set for collective funding? Clearly, the answer will depend on the outcomes of further debate and research on WGS-PGT’s clinical utility and proportionality.
